# SchistoTrackNet: machine learning for diagnosis of schistosomiasis-associated periportal fibrosis from ultrasound images

**DOI:** 10.64898/2026.06.01.26354609

**Authors:** Eloise Ockenden, Victor Anguajibi, Simon Mpooya, Benjamin Ntegeka, Tymothy Mugume, Betty Nabatte, Narcis Kabatereine, J. Alison Noble, Goylette F. Chami

**Affiliations:** Nuffield Department of Population Health, University of Oxford, Oxford, United Kingdom; Institute of Biomedical Engineering, University of Oxford, Oxford, United Kingdom; Division of Vector-Borne and Neglected Tropical Diseases Control, Uganda Ministry of Health, 15 Bombo Road, Kampala, Uganda

## Abstract

Liver fibrosis is a major cause of death in low- and middle-income country contexts. In rural, poor areas of sub-Saharan Africa, schistosomiasis is an underestimated cause of liver fibrosis. Despite the need for increased diagnostic capacity for schistosomiasis-related liver fibrosis, there are no automated, clinically-validated tools to diagnose schistosomiasis-related liver fibrosis. We present SchistoTrackNet which is, to our knowledge, the first deep learning-based model for distinguishing distinct presentations of schistosomiasis-related liver fibrosis of varying severity. Ultrasound images from 1533 participants aged 5–84 years from three districts in rural Uganda were used to train and evaluate the presented models. The models were evaluated by assessing failure cases and by comparing results with re-readings performed by sonographers experienced in diagnosis of schistosomiasis morbidity. Our models show potential to enable automated reading of ultrasound images for schistosomiasis-related liver fibrosis to allow large-scale surveillance of schistosomiasis morbidity and contribute towards the World Health Organization target to eliminate schistosomiasis as a public health problem.

## INTRODUCTION

Liver fibrosis is a leading cause of death and disability worldwide, often caused by complex infections in low- and middle-income countries (LMICs) [1]. Intestinal schistosomiasis, in particular that caused by *Schistosoma mansoni*, is a historically underestimated cause of liver fibrosis in sub-Saharan Africa (SSA) [2]. Over 700 million people in SSA are estimated to be at risk of schistosomiasis infection, which is transmitted by a water-borne parasitic blood fluke [3]. The World Health Organization (WHO) roadmap for neglected tropical diseases (NTDs) [4] targets the elimination of schistosomiasis as a public health problem by 2030, which requires elimination of severe disease. However, WHO guidelines focus on current infection intensity as a proxy indicator of morbidity, aiming to reduce the percentage of people with heavy infections to below 1% of the infected population [4, 5]. Recent work in Uganda and a meta-analysis of 33 studies have shown no correlation of current intestinal schistosome infection with liver fibrosis [6, 7]. Consequently, World Health Organization Regional Office for Africa (WHO-AFRO) and other international expert groups have called for better morbidity indicators, which could meet gaps in expertise and diagnostic resources in schistosomiasis-endemic countries [8]. To our knowledge, there are no automated and clinically-validated ultrasound image analysis tools for differential morbidity diagnosis related to schistosomiasis.

Intestinal schistosomiasis can result in diverse forms of liver fibrosis, broadly termed periportal fibrosis (PPF). Eggs laid by female flukes, residing in the mesenteric veins, may get carried through the portal blood flow to pre-sinusoidal vessels [9]. Granulomatous, inflammation-driven immune responses to antigens excreted by the eggs can, if uncontrolled, lead to liver fibrosis. If the peripheral portal vasculature is impaired, intra-hepatic portal pressure increases, which allows eggs to be trapped in larger (lower order) portal branches [9]. Periportal fibrosis (PPF) can range from localised changes in the periportal space to extensive fibrosis extending to the liver capsule. The diverse presentations of PPF result in a restructuring of the liver vasculature and distinctive patterns of fibrosis that can be captured and visualised using B-mode ultrasound imaging. Point-of-care ultrasound imaging (POCUS) is the most-used diagnostic tool for schistosomal PPF due to its relatively low cost and portability. The most recent WHO guidance for diagnosis and staging of PPF is the Niamey protocol [10, 11]. Despite the lack of sonography training and expertise in LMICs [12], use of the Niamey protocol requires extensive expertise and exposure to many cases of diverse presentations of liver fibrosis. Fully automated deep learning-based models have yet to be developed to support diagnoses of liver fibrosis patterns described by the Niamey protocol, and it remains an open question as to whether such models could perform comparably with second sonographer re-readings of ultrasound images.

Moreover, for research studies and clinical trials, deep learning-based models for ultrasound imaging of the liver could build capacity for re-readings or second readers to validate diagnoses, needed due to the subjective nature of reading medical images. To our knowledge, there are no studies that have used deep learning-based models as a second reader of ultrasound images of liver fibrosis, and for PPF, very few studies have even used a second (human) reader to validate diagnoses [11]. Diagnoses of PPF in schistosomiasis-related clinical trials is typically done using second readers who provide the final definitive, and only, diagnosis using images acquired by another sonographer where the second reader has no involvement in examining the patient [13]. A limitation of this approach is that diagnoses may be unconfirmed, and the sonographer can choose the view of the liver and pathology to focus on when making a diagnosis, which is not accounted for when only reading images separately from acquisition. Therefore, there is a current need for studies that validate deep learning-based models as second readers by comparing their performance against human readers.

Here we developed, to our knowledge, the first deep learning-based models for fully automated classification of differential patterns of schistosomal PPF and related liver fibrosis. We compared convolutional neural network (CNN)-based and transformer-based models. Models were built and evaluated using B-mode point-of-care ultrasound imaging (POCUS) from 1533 participants with and without liver fibrosis, acquired by four sonographers between January–February 2023 and January–February 2024, as part of the SchistoTrack cohort study. Participants were aged 5–84 years and came from three districts in rural Uganda. The aim of our study was to assess whether deep learning-based models can differentially diagnose diverse types of schistosomal PPF, perform comparably with expert sonographers, and serve as second readers of liver ultrasound images.

## RESULTS

### Liver fibrosis prevalence

There were 871 participants from 2023 and 1039 participants from 2024. A total of 1533 participants were studied across the two years, with 377 participants contributing in both years. For the 2023 data, participants were aged 5–84 years at the time of sonography assessment, with a mean age of 29.3 (standard deviation (s. d.) 19.0) and 45.9% (400/871) were female. For the 2024 data, participants were aged 5–82 years at the time of sonography assessment, with a mean age of 30.7 (s. d. 19.2) and 48.7% (506/1039) were female. For the 2023 data, 53.5% (466/871) participants were from Pakwach, 30.2% (263/871) were from Buliisa and 16.3% (142/871) were from Mayuge. For the 2024 data, 53.0% (551/1039) participants were from Pakwach, 29.0% (301/1039) were from Buliisa and 18.0% (187/1039) were from Mayuge.

Liver fibrosis patterns were represented by letters B–F, where C–F patterns were termed periportal fibrosis (PPF). Among the 871 participants from 2023, 100 had no observable liver fibrosis patterns and the remaining 771 had at least one liver fibrosis pattern. All 1039 participants from 2024 had at least one liver fibrosis pattern. 55.5% (483/871) of included participants had PPF at the 2023 time-point, and 56.1% (583/1039) had PPF at the 2024 time-point. Among participants from the 2023 time-point, 44.0% (393/871) had B liver fibrosis patterns, representing diffuse liver fibrosis patterns not necessarily attributable to schistosomiasis. Among participants from the 2024 time-point, 56.7% (589/1039) had B liver fibrosis patterns. The numbers of participants at each time-point with patterns C1 or C2, representing mild fibrosis concentrated on the portal vessels and pre-sinusoidal vessels; pattern D, representing moderate fibrosis adjacent to the main portal vein; and patterns E or F, representing severe liver fibrosis extending to the liver capsule and including blockage of the main portal vein are shown in Supplementary Table S3. The distribution of the liver fibrosis patterns among adults (defined as participants aged 15 years or above) and children is given in Supplementary Table S4.

A total of 3710 ultrasound images of curated views of liver fibrosis patterns were collected from 771 and 1039 participants in 2023 and 2024 respectively, with any type of liver fibrosis. 1433 participants contributed images of liver fibrosis patterns across the two years. Each image had one fibrosis pattern label. The median number of images per participant, among individuals with fibrosis, was two (interquartile range (IQR) 1– 3). The overall distribution of fibrosis images is shown in Supplementary Table S3. In addition to the images of fibrosis patterns, 100 participants without any liver fibrosis patterns were selected from the 2023 time-point and four frames were taken from liver ultrasound video of each of these participants to form a healthy class of 400 images.

### Performance of deep learning models for liver fibrosis pattern classification

We compared the performance of five deep learning-based models: two CNN-based models, SchistoTrackNet and SchistoTrackNet-Random; and three transformer-based models using the vision transformer (ViT) [14], ViT-SupCon (supervised contrastive loss), ViT-Focal and ViT-CE (cross entropy loss) (Table 1). SchistoTrackNet used a PULSENet encoder for initialisation before refining the encoder on liver ultrasound image data from SchistoTrack. The variant of PULSENet that was used was an encoder trained to classify 13 fetal anatomies from first trimester ultrasound images from the PULSE dataset [15, 16]. Thus the encoder was familiar with ultrasound image appearance but not the specific ultrasound application and anatomy. SchistoTrackNet-Random had the same architecture but was initialised randomly. The ViT-based models were initialised using ImageNet weights [17]. To evaluate the 10-fold cross validation results, Friedman’s *χ*^2^-test was performed on the F1-score, giving a *χ*^2^ statistic of 20.96, *p* = 0.0003, indicating statistically significant differences between model performances. Therefore, post-hoc Wilcoxon signed-rank tests were run between all pairs of models, with a Holm correction to account for multiple testing. Overall, differences were minimal with only two pairs of models having statistically significant differences in F1-score. The significant differences in F1-score were between the SchistoTrackNet and ViT-Focal (*p* = 0.0195), and between SchistoTrackNet and ViT-CE (*p* = 0.0176).

**Table 1:** Overall model performance. A summary of the quantitative results gained from the different models that were used. For the results given using 10-fold cross validation, the mean value of the metric across the 10 folds is given, and the standard deviation is given in brackets. For each metric, for the cross validation and for the test set, bootstrapped 95% confidence intervals are shown in square brackets.

| Model | Accuracy | Balanced accuracy | F1-score |
| --- | --- | --- | --- |
| 10-fold cross validation |  |  |  |
| *CNN-based* |  |  |  |
| SchistoTrackNet | **0.8516** (0.016)<br>[0.843, 0.861] | **0.7832** (0.039)<br>[0.760, 0.806] | **0.8497** (0.016)<br>[0.841, 0.860] |
| SchistoTrackNet-Random | 0.8337 (0.027)<br>[0.818, 0.849] | 0.7534 (0.054)<br>[0.721, 0.784] | 0.8319 (0.029)<br>[0.813, 0.846] |
| *Transformer-based* |  |  |  |
| ViT-SupCon | 0.8318 (0.021)<br>[0.820, 0.844] | 0.7563 (0.025)<br>[0.742, 0.771] | 0.8287 (0.022)<br>[0.815, 0.842] |
| ViT-Focal | 0.8158 (0.020)<br>[0.805, 0.827] | 0.7386 (0.024)<br>[0.725, 0.753] | 0.8137 (0.020)<br>[0.802, 0.825] |
| ViT-CE | 0.8209 (0.015)<br>[0.812, 0.830] | 0.7514 (0.024)<br>[0.737, 0.766] | 0.8187 (0.015)<br>[0.810, 0.828] |
| Test set |  |  |  |
| *CNN-based* |  |  |  |
| SchistoTrackNet | **0.8222**<br>[0.785, 0.857] | **0.8121**<br>[0.763, 0.861] | **0.8223**<br>[0.784, 0.858] |
| SchistoTrackNet-Random | 0.8037<br>[0.764, 0.838] | 0.6972<br>[0.665, 0.729] | 0.7877<br>[0.747, 0.826] |
| *Transformer-based* |  |  |  |
| ViT-SupCon | 0.8014<br>[0.762, 0.838] | 0.7300<br>[0.681, 0.776] | 0.7944<br>[0.753, 0.833] |
| ViT-Focal | 0.7852<br>[0.748, 0.824] | 0.7130<br>[0.659, 0.764] | 0.7771<br>[0.736, 0.818] |
| ViT-CE | 0.7783<br>[0.739, 0.818] | 0.7282<br>[0.678, 0.782] | 0.7739<br>[0.733, 0.814] |

On the test set, all models achieved high performance when compared to the sonographer who acquired images in the field. SchistoTrackNet had the highest accuracy (0.8222), balanced accuracy (0.8121), and F1-score (0.8223) of any of the models. When comparing the bootstrapped 95% confidence intervals between SchistoTrackNet and the other models, there was crossover between all confidence intervals for accuracy and F1-score; indicating that differences were not necessarily statistically significantly different. SchistoTrackNet achieved the highest balanced accuracy by eight percentage points. However, the only statistically significant difference in balanced accuracy was between SchistoTrackNet and SchistoTrackNet-Random. Uniform manifold approximation and projection (UMAP) projections of the feature vectors given by each model are shown in Supplementary Figure S2, showing clear clusters for each of the six classes for all models aside from SchistoTrackNet-Random.

Figure 1 shows confusion matrices for liver patterns as compared to the diagnoses given by expert sonographers at the point of participant examination. All models performed well in classifying images of healthy livers with sensitivities ranging from 87.5% for the ViT-Focal to 97.5% for ViT-SupCon and SchistoTrackNet-Random. SchistoTrackNet had the most consistent performance across PPF patterns; obtaining the highest percentage of correct classifications for mild, moderate and severe PPF patterns (C2, D and E/F), which were 84.5%, 91.4% and 57.9% respectively. The B pattern (which does not necessarily relate to schistosomiasis) and the C1 pattern, which represents the same severity but a different view of the C2 pattern, were best classified by ViT-Focal. Across the five models, the liver patterns that are found in the transverse view (B, C2 and E/F) were likely to be confused with one another. In particular, for each of the five models, true B pattern images were most likely to be predicted as a C2 pattern when misclassified, and vice versa. Similarly, aside from for SchistoTrackNet, true E/F patterns were most likely to be misclassified as the C2 pattern. Cohen’s *κ* was the highest for SchistoTrackNet, at 0.772. The range of Cohen’s *κ* values was 0.712–0.772 across the five models.

**Fig. 1:**
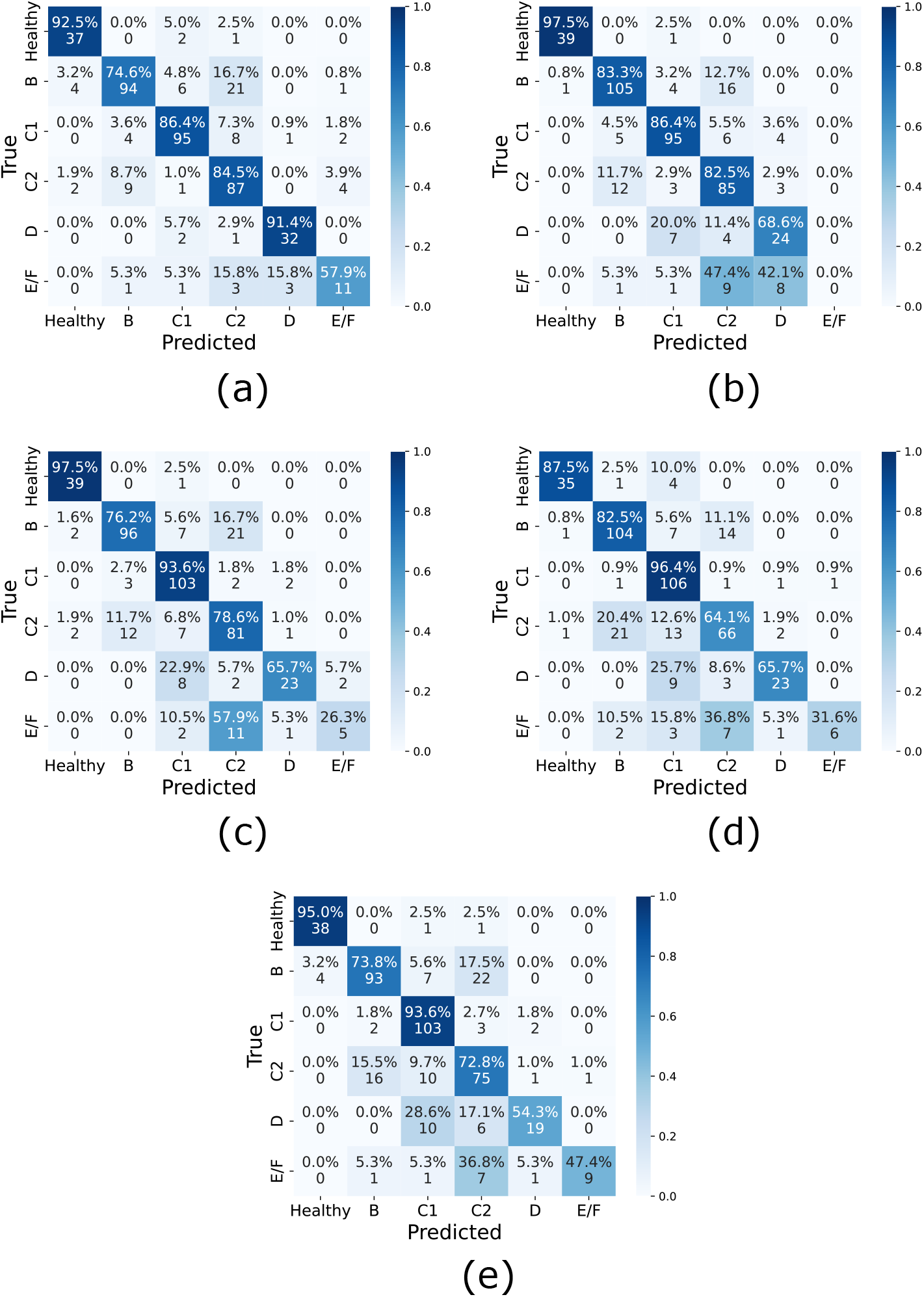
Confusion matrices for deep learning-based models. (a) SchistoTrack-Net. (b) SchistoTrackNet-Random. (c) ViT-SupCon. (d) ViT-Focal. (e) ViT-CE.

To understand the clinical relevance of the model predictions, Table 2 shows the positive predictive values (PPVs) and negative predictive values (NPVs) for each model on the test set. The NPV was high across all liver patterns for all models, in most cases being *>* 0.90. The PPV was variable among the liver patterns, reflecting the variable sensitivity across the patterns. SchistoTrackNet had the bestPPVs of all models for the B, C1 and D patterns, and the best NPVs in the C2, D and E/F patterns. In particular, for the D pattern, SchistoTrackNet had high sensitivity (0.9143) and a high PPV (0.8889), showing that cases that were detected were very likely to be true cases of D patterns, according to the pattern assigned at the point of acquisition.

**Table 2:** Positive and negative predictive values by liver fibrosis pattern. A summary of the PPV and NPV achieved by each model with respect to the diagnoses given at the point of image acquisition. The highest PPV and NPV across the five models for each liver fibrosis pattern is emboldened.

| Model | Healthy | B | C1 | C2 | D | E/F |
| --- | --- | --- | --- | --- | --- | --- |
| *CNN-based* |  |  |  |  |  |  |
| *SchistoTrackNet* |  |  |  |  |  |  |
| PPV | 0.8605 | **0.8704** | **0.8879** | 0.7190 | **0.8889** | 0.6111 |
| NPV | 0.9923 | 0.9015 | 0.9540 | **0.9487** | **0.9924** | **0.9807** |
| Sensitivity | 0.9250 | 0.7460 | 0.8636 | 0.8447 | 0.9143 | 0.5789 |
| Specificity | 0.9847 | 0.9544 | 0.9628 | 0.8970 | 0.9899 | 0.9831 |
| *SchistoTrackNet-Random* |  |  |  |  |  |  |
| PPV | **0.9750** | 0.8537 | 0.8559 | 0.7083 | 0.6154 | 0.0000 |
| NPV | 0.9975 | **0.9323** | 0.9534 | 0.9425 | 0.9721 | 0.9561 |
| Sensitivity | 0.9750 | 0.8333 | 0.8636 | 0.8252 | 0.6857 | 0.0000 |
| Specificity | 0.9975 | 0.9414 | 0.9505 | 0.8939 | 0.9623 | 1.0000 |
| *Transformer-based* |  |  |  |  |  |  |
| *ViT-SupCon* |  |  |  |  |  |  |
| PPV | 0.9070 | 0.8649 | 0.8047 | 0.6923 | 0.8519 | 0.7143 |
| NPV | 0.9974 | 0.9068 | 0.9770 | 0.9304 | 0.9704 | 0.9671 |
| Sensitivity | 0.9750 | 0.7619 | 0.9364 | 0.7864 | 0.6571 | 0.2632 |
| Specificity | 0.9898 | 0.9511 | 0.9226 | 0.8909 | 0.9899 | 0.9952 |
| *ViT-Focal* |  |  |  |  |  |  |
| PPV | 0.9459 | 0.8062 | 0.7465 | **0.7253** | 0.8519 | 0.8571 |
| NPV | 0.9874 | 0.9276 | **0.9863** | 0.8918 | 0.9704 | 0.9695 |
| Sensitivity | 0.8750 | 0.8254 | 0.9636 | 0.6408 | 0.6571 | 0.3158 |
| Specificity | 0.9949 | 0.9186 | 0.8885 | 0.9242 | 0.9899 | 0.9976 |
| *ViT-CE* |  |  |  |  |  |  |
| PPV | 0.9048 | 0.8304 | 0.7803 | 0.6579 | 0.8261 | **0.9000** |
| NPV | **0.9949** | 0.8972 | 0.9767 | 0.9122 | 0.9610 | 0.9764 |
| Sensitivity | 0.9500 | 0.7381 | 0.9364 | 0.7282 | 0.5429 | 0.4737 |
| Specificity | 0.9898 | 0.9381 | 0.9102 | 0.8818 | 0.9899 | 0.9976 |

We investigated images that had been predicted correctly versus incorrectly, to understand the types of errors that the models were making. Figure 2(a) shows an image that was classified correctly as an E/F grade for three models (SchistoTrackNet, ViT-SupCon and ViT-Focal), whereas Figure 2(b) shows an image that was assigned as E/F at the field scan, but was assigned C2 by the same three models. The portal vein is visible in this image, which was confused with portal vein branches in a true C2 image (Figure 2(d)). For C-pattern images, Figure 2(c) shows a C1-pattern image characterised by the ‘rings’ that appear due to the fibrosis in the portal vein branches. Figure 2(e) shows an example of an image that was assigned C1 at the field scan, but was assigned C2 by SchistoTrackNet, ViT-SupCon and ViT-Focal. This image shows portal vein branches coming across the field of view, which is a key anatomical characteristic of C2 images.

**Fig. 2:**
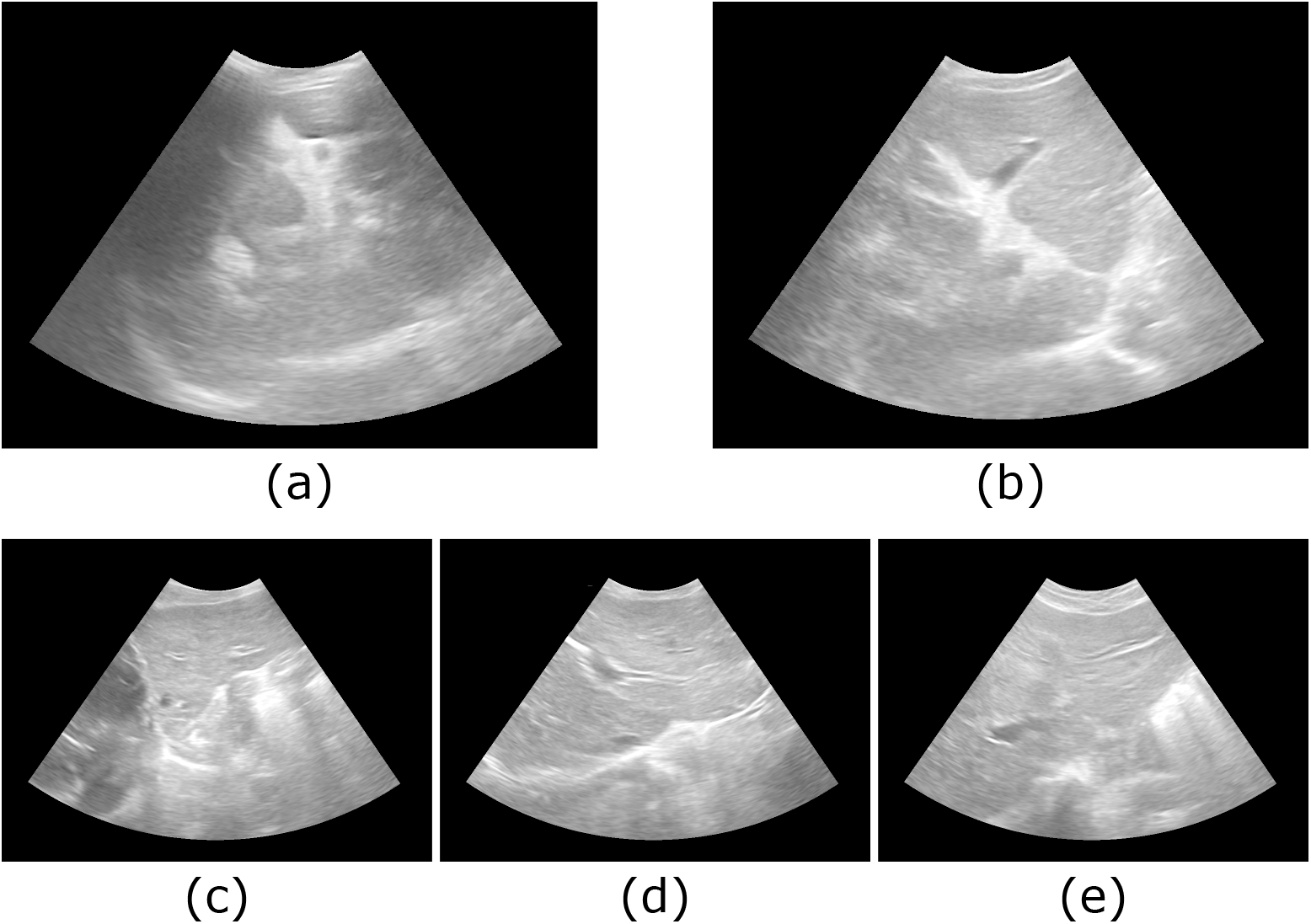
Example ultrasound images of C and E/F patterns. (a) E/F pattern image correctly predicted. (b) E/F pattern image predicted as C2. (c) C1 pattern image correctly predicted. (d) C2 pattern image correctly predicted. (e) C1 pattern image predicted as C2.

Since images of healthy livers were not collected and video frames were used instead, we present results for models trained without healthy participants in Supplementary Figure S10 and Table S7. The results using a model that excludes a healthy class were similar to results including a healthy class, although the overall accuracy of a model was higher when the healthy class was included. We hypothesise that this is because it is an easier task to distinguish healthy video frames from images of diseased livers.

### Machine learning models as second readers

We measured agreement between a human sonographer making a diagnosis at the point of patient examination and ultrasound image acquisition (here called the field sonographer), and a sonographer re-reading an image after ultrasound data collection with a laptop (here called the second sonographer) (Figure 3(a)). The second sonographer only had access to the unlabelled ultrasound image and no other context about the image or participant. There was low agreement between the field and second sonographers for the mild fibrosis patterns, particularly for the B pattern where agreement was 46.0%. The B pattern was most often read by the second reader as healthy or as a C1 pattern. The C1 and C2 patterns, which showed distinct views of the same severity of fibrosis, had the highest agreement at 73.6% to 74.9%. The moderate and severe liver patterns (D and E/F) showed similar agreement, at around 68%. Overall, Cohen’s *κ* for the agreement between the field and second sonographers was 0.54, compared with 0.77 for SchistoTrackNet compared to the field sonographer. The percentage of images agreed upon between SchistoTrackNet and the field sonographer was higher than the second sonographer for every pattern aside from the most severe pattern (E/F). These differences remained when analysed separately by year (shown in Supplementary Figures S3, S4 and S5). SchistoTrackNet classified more images correctly than agreed upon by the second sonographer for all liver patterns aside from C2 for 2023 data. For 2024 data, SchistoTrackNet classified more images correctly than agreed upon by the second sonographer for all liver patterns aside from the E/F pattern.

**Fig. 3:**
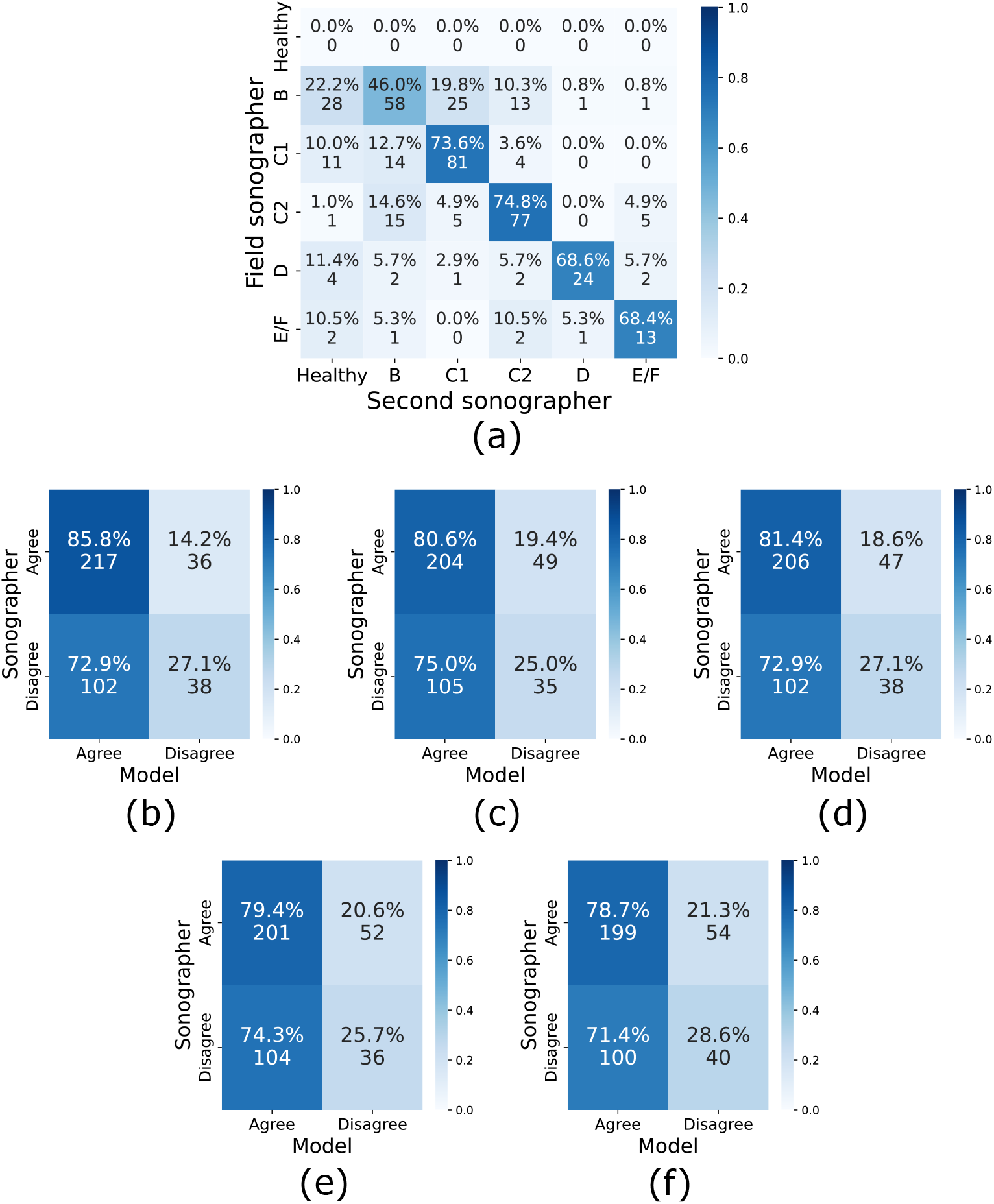
Sonographer agreement. (a) This confusion matrix compares the liver pattern assigned at the point of image acquisition (field scan) and the liver pattern assigned by the second sonographer reader, for the test set of curated images. (b)–(f) These confusion matrices show the number of images that were agreed upon by the sonographers and by the model, for the test set. (b) SchistoTrackNet. (c) SchistoTrackNet-Random. (d) ViT-SupCon. (e) ViT-Focal. (f) ViT-CE.

There was some overlap between images disagreed upon by each of the models and images disagreed upon by the second sonographer, shown in Figure 3(b)–(f). Images that were disagreed upon between the field and second sonographers were more likely to be misclassified by SchistoTrackNet and all other models, than those that were agreed upon. Similarly to the confusion of the patterns by the five models, the second sonographer also confused patterns in the transverse view (the B, C2 and E/F patterns). For true E/F pattern images, the second sonographer was most likely to read them as C2 or healthy; and for true C2 pattern images the second sonographer was most likely to read them as a B pattern. However, true B pattern images were most likely to be read as healthy.

SchistoTrackNet focused on areas of the liver with fibrosis for some patterns. Figure 4 shows activation maps given by GRAD-CAM [18, 19] alongside ultrasound images annotated by experienced sonographers. The D pattern images (Figures 4(h), (i) and (j)) were the most consistent in having high activation in the area of the image where the D pattern of fibrosis occurs, around the main portal vein. Some images of other patterns (B2, C1, C2, and E/F) shown in Figures 4(b), (d), (g), and (k) also showed areas of high activation where liver patterns occurred. However, areas of high activation could also be observed where there were other anatomical features such as the diaphragm (Figure 4(f)), or interference such as gas (Figures 4(a) and (e)). Attention maps were also produced using attention rollout [20] from the ViT-SupCon (Supplementary Figure S11). Attention was more likely to be distributed throughout the field of view than the activation and often was focused on areas with gas interference.

**Fig. 4:**
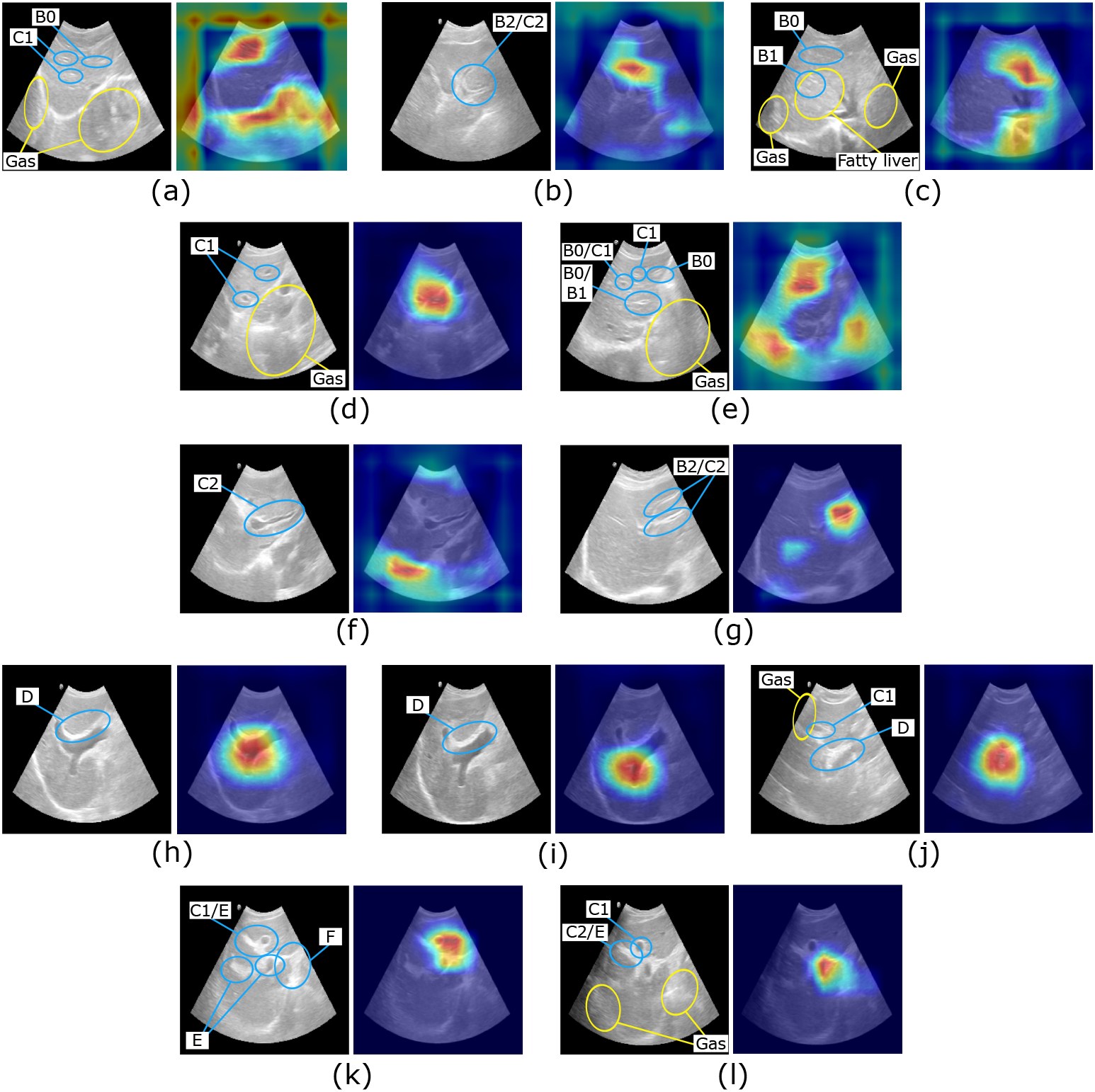
Annotated ultrasound images with activation maps. Ultrasound images for each grade and with different predictions (correctly or incorrectly predicted by SchistoTrackNet) alongside activation maps given by GRAD-CAM from SchistoTrack-Net. Red areas of the image show high activation and blue areas show low activation.(a) B0, correctly predicted. (b) B2, incorrectly predicted (E/F). (c) B1b, correctly predicted. (d) C1, correctly predicted. (e) C1, incorrectly predicted (C2). (f) C2, correctly predicted. (g) C2, incorrectly predicted (healthy). (h) D, correctly predicted. (i) D, correctly predicted. (j) D, correctly predicted. (k) E, correctly predicted. (l) E, incorrectly predicted (C2).

### Lack of influence of patient-level factors on model performance

Patient-level factors, such as age, sex, fasting status, pregnancy and common comorbidities had little effect on model performance for identifying patterns of fibrosis (Table 3). These factors may cause difficulty identifying schistosomiasis-associated liver fibrosis patterns. Not fasting may cause abdominal gas which can negatively affect image quality; hepatitis B may result in liver damage resulting in missed fibrosis patterns; and fatty livers are highly echogenic causing additional brightness in the liver which may obscure fibrosis patterns. *χ*^2^ tests showed that the proportion of errors was not significantly different for any model for individuals who did not fast or had hepatitis B, fatty livers, or were pregnant at the time of examination. For age and sex, there were no significant differences except for adults when SchistoTrackNet was used. This difference may be driven by the lower sensitivity for the B patterns (57%) in adults for this model (shown in Supplementary Figure S6). Notably, B patterns were less prevalent in adults.

**Table 3:** Patient-level factors on model performance on the test set. A summary of the numbers of errors in adults (defined as participants aged ≥ 15, female participants, participants with the most common sonography findings and participants who did not fast for at least 2 hours before the ultrasound examination, for the test set.

| Model | Proportion<br>among images | Number of<br>errors | $\chi^2$ (p-value) |
| --- | --- | --- | --- |
| *CNN-based* |  |  |  |
| *SchistoTrackNet* |  |  |  |
| Adults | 77.1% (303/393) | 66/74 | 6.12 (0.01)* |
| Female | 53.6% (211/393) | 46/74 | 2.14 (0.14) |
| Not fasting | 15.5% (61/393) | 12/74 | 0.03 (0.87) |
| Hepatitis B | 1.5% (6/393) | 1/74 | 0.02 (0.90) |
| Fatty liver | 0.8% (3/393) | 1/74 | 0.34 (0.56) |
| Pregnancy | 0.8% (3/393) | 0/74 | 0.57 (0.45) |
| *SchistoTrackNet-Random* |  |  |  |
| Adults | 77.1% (303/393) | 72/84 | 3.53 (0.06) |
| Female | 53.6% (211/393) | 44/84 | 0.06 (0.81) |
| Not fasting | 15.5% (61/393) | 14/84 | 0.08 (0.77) |
| Hepatitis B | 1.5% (6/393) | 1/84 | 0.06 (0.80) |
| Fatty liver | 0.8% (3/393) | 1/84 | 0.20 (0.65) |
| Pregnancy | 0.8% (3/393) | 1/84 | 0.20 (0.65) |
| *Transformer-based* |  |  |  |
| *ViT-SupCon* |  |  |  |
| Adults | 77.1% (303/393) | 72/85 | 2.79 (0.10) |
| Female | 53.6% (211/393) | 37/85 | 3.53 (0.06) |
| Not fasting | 15.5% (61/393) | 16/85 | 0.71 (0.40) |
| Hepatitis B | 1.5% (6/393) | 2/85 | 0.39 (0.53) |
| Fatty liver | 0.8% (3/393) | 2/85 | 2.84 (0.09) |
| Pregnancy | 0.8% (3/393) | 0/85 | 0.65 (0.42) |
| *ViT-Focal* |  |  |  |
| Adults | 77.1% (303/393) | 75/88 | 3.29 (0.07) |
| Female | 53.6% (211/393) | 46/88 | 0.07 (0.79) |
| Not fasting | 15.5% (61/393) | 18/88 | 1.63 (0.20) |
| Hepatitis B | 1.5% (6/393) | 1/88 | 0.09 (0.77) |
| Fatty liver | 0.8% (3/393) | 2/88 | 2.65 (0.10) |
| Pregnancy | 0.8% (3/393) | 0/88 | 0.68 (0.41) |
| *ViT-CE* |  |  |  |
| Adults | 77.1% (303/393) | 80/94 | 3.41 (0.06) |
| Female | 53.6% (211/393) | 41/94 | 3.84 (0.05) |
| Not fasting | 15.5% (61/393) | 15/94 | 0.01 (0.91) |
| Hepatitis B | 1.5% (6/393) | 2/94 | 0.23 (0.63) |
| Fatty liver | 0.8% (3/393) | 1/94 | 0.11 (0.74) |
| Pregnancy | 0.8% (3/393) | 0/94 | 0.72 (0.40) |

## DISCUSSION

The WHO has targeted schistosomiasis for elimination as a public health problem by 2030 in its 2021 roadmap for NTDs [4]. However, this target focuses on eliminating heavy infections rather than liver morbidity. To reach the target of elimination as a public health problem, better morbidity estimates for schistosomiasis are needed, in particular for schistosomal liver fibrosis which requires new tools for diagnosis. In this study, 1533 participants aged 5–84 from three districts in rural Uganda were examined using B-mode ultrasound imaging. From these participants, images were taken of liver patterns indicative of schistosomal PPF. We developed the first deep learning-based models for distinguishing the distinct patterns of fibrosis associated with PPF. The presented models classify these complex patterns of fibrosis with high accuracy (up to 82.2%) and perform comparably to re-reading of images by second sonographers, despite the presence of comorbidities, differences in fasting, pregnancy and the diverse demographics of the population.

All models that are presented in this paper performed comparably with expert sonographers with 8–20 years of experience diagnosing schistosomiasis-related morbidity. Our best-performing model—SchistoTrackNet—yielded an accuracy of 82.2% on an unseen test set. There is limited previous literature analysing ultrasound images of schistosomiasis-induced liver fibrosis; there is one study on *Schistosoma japonicum* in China [21] which analysed ultrasound images of schistosomal PPF patterns using radiomics features typically used in cancer image analyses. Results were presented as binary comparisons between each pair of patterns with focus on comparing adjacent patterns, with areas under the receiver operating curve (AUCs) ranging from 0.771–0.834. To our knowledge, there is only one study that has used deep learning-based methods on ultrasound images of schistosomal liver fibrosis [22]. In that study, a CNN-based architecture for a binary problem; to distinguish between livers with and without PPF, achieving an accuracy of 82% and F1-score of 0.82. 160 images were used for training this model, and 40 were used for testing. To our knowledge, there are no models that distinguish the distinct types of schistosomal PPF. Our models performed well on a six-class problem using, to our knowledge, the largest available dataset for schistosomiasis-related morbidity in SSA. Our models were able to distinguish between mild and advanced liver fibrosis. Of the five models we considered, SchistoTrackNet was the best model for dealing with class imbalance present in the dataset, achieving a balanced accuracy of 0.8121 on an unseen test set. We conclude from this that SchistoTrackNet is the most promising for further validation in other contexts and for the development of a formal tool for re-reading of liver ultrasound images. However, for some classes, such the C1 liver pattern and healthy livers, ViT-SupCon was found to be more sensitive. Future work could consider integrating data from other countries/regions, and develop lightweight models that can be run on portable tablets before roll out of these models for patient diagnoses within health facilities or morbidity surveillance in national control programmes.

We presented the sensitivity and specificity achieved by our models for identification of liver fibrosis patterns, which exceeded those attained by the second sonographer for most patterns. The specificity was high for all image patterns, surpassing 90% in every pattern aside from the C2 pattern, where specificity was 89.7%. The sensitivity for the D image pattern, representing ruffing around the main portal vein, was *>* 90%, and was also high for the C1 and C2 patterns (86.4% and 84.5% respectively). In the early stages of monitoring and evaluating schistosomiasis control programmes, specificity should be prioritised for diagnostic tests with a WHO target product profile for diagnosis of infection suggesting a minimum specificity threshold of 95%, by contrast to a minimum sensitivity threshold of 60% [23]. In the clinic, high sensitivity for severe liver patterns that may require urgent care is important so that appropriate care can be provided. In a related problem for a different field, a breast cancer meta-analysis showed that machine learning-based tools had higher sensitivity but lower specificity than radiologists [24]. However, in low-resource clinical settings, it is important that specificity is retained, since resources are scarce and cannot be wasted on false positives. The PPVs and NPVs for SchistoTrackNet were high compared to other deep learning-based models for medical image re-reading [25]. For the D liver pattern, for which the sensitivity was 91.4%, the PPV was 0.8889. Additional data and clinical trials of our model are needed for implementation as a diagnostic tool. In addition to this, workshops and demonstrations across endemic countries would be needed to identify issues regarding implementation, trust and acceptance for users of the tool in their routine patient flows.

We have presented models with clinically explainable misclassifications and provide evidence for needed updates to WHO protocol guidance for the diagnosis of schistosomiasis morbidity, which is now outdated since it was last updated in 2000 [10]. Several of the liver patterns (B, C2 and E/F patterns) are found in the same, transverse view of the liver, and these were most often confused with each other, rather than patterns being confused due to similar severity. The confusion between B, C2 and E/F patterns may be evidence of the model learning the transverse view of the liver rather than the severity of the morbidity, which reflects the way that sonographers learn to diagnose liver patterns in training. At re-reading, the second sonographer was also more likely to confuse patterns in the same (transverse) view. The views of the liver detailed as part of the SchistoTrack data collection protocol should be considered for incorporation into future versions of the Niamey protocol to enable better reproducibility and standardisation. The standardised views may also be useful for creating an image bank for the WHO to accompany the Niamey protocol [10]. Use of the Niamey protocol relies upon exposure to many cases of liver patterns, which is not currently integrated into sonographer training in schistosomiasis-endemic countries. Provision of example images from this study could provide a training aid for new sonographers to increase capacity for PPF diagnosis for clinical and research settings. Secondly, the Niamey protocol advises against diagnosis in cases of fatty liver, but this exclusion that may result in many people remaining undiagnosed and untreated may be unnecessary. Our findings show that images from participants with comorbidities such as fatty liver, and who did not fast were not more likely to be misclassified by the model than other participants. Capturing the continuous nature of fibrosis progression needs to be explored in further work, since the Niamey protocol only presents discrete categories of fibrosis. Overall, an update to the Niamey protocol is needed that incorporates image banks, explicitly describes training tools, and better considers comorbidities when diagnosing PPF.

There is potential for deep learning-based models to serve as second readers of ultrasound images for schistosomal PPF diagnosis, which could be useful both for clinical settings and clinical trials [26, 27]. In this paper, we used the diagnosis given in the field as the ground truth because more information may be contained in the labels given, such as patient characteristics and other context from doing preliminary scans; and because it avoids laboratory bias [28]. SchistoTrackNet agreed with the diagnosis given at the point of image acquisition more often than the second sonographer, despite the model being given the same information as the second sonographer: an ultrasound image only with no other context about the patient. The improved agreement between SchistoTrackNet and the diagnosis given at image acquisition compared to the second sonographer persists despite giving the second sonographer an advantage. The second sonographer in this study was able to assign multiple liver patterns for a single ultrasound image, whereas all models here were only given one shot at the correct classification. Further work is needed to explore how a deep learning-based model could be used as second reader for schistosomal PPF. Possible scenarios, as have been explored in breast cancer imaging [29], are for one radiologist to read an image and for a deep learning-based model to re-read, or for a deep learning-based model to select images for human re-reading where only images that have disagreement between the first reader and the model are sent for a second human reading.

Key strengths in this study lie with the large scale of the study of schistosomiasis, examination of a population with high disease burden, high performing deep learning-based models, and validation of models through clinical review of model performance. Despite this, our study has several limitations. There were no images collected from healthy participants to create a healthy class. Therefore, we used video frames from participants without patterns of liver fibrosis. A further limitation is that the models presented in this paper were only trained and tested on data collected by one type of ultrasound machine and using data from one country. Future work should explore the application of these models to data collected using cheaper, lower-quality portable ultrasound as well as for individuals from other countries.

Here, we present the first deep learning-based models able to differentiate patterns of schistosomal liver fibrosis. SchistoTrackNet had a high diagnostic accuracy and gave diagnoses that agreed with the field scan more than a second sonographer at re-reading. Additional work is needed to incorporate data from settings outside of Uganda, to run clinical trials to assess how the use of deep learning-based models may improve patient diagnoses and long-term clinical outcomes, as well as save resources and time for limited capacity settings. We anticipate that the proposed model may serve as a starting point for tools for large-scale surveillance of schistosomiasis morbidity in endemic settings, promote the training of additional sonographers in resource-limited settings and enable updates to the global burden of schistosomiasis-related disease to better target resources to patients and geographic areas most in need of improved care.

## METHODS

### Study design

The SchistoTrack cohort is a prospective, longitudinal community-based study of individuals. The first year of observation was conducted in 2022 and there is annual follow-up planned until 2027 [30, 31]. The cohort is based in three districts of rural Uganda: Pakwach, Buliisa and Mayuge. Two of the districts, Pakwach and Buliisa, are in the west of Uganda, bordering the White Nile and Lake Albert respectively. Buliisa and Mayuge both have hospitals in the district, however the highest level of health centre that Pakwach has is a Health Centre IV. Households were sampled for participation in the study from 52 villages across the three districts using a local register. Uniform random sampling was used to sample 40 households from each register, with oversampling of 30 households per village to allow for ineligible households, refusal or other reasons for non-participation. One child (aged 5–17 years) and one adult (aged ≥ 18 years) were put forward from each household by the household head or their spouse for clinical participation in the study. This study focuses on participants that participated in clinical data collection in 2023 and 2024. More information on the study cohort can be found here [30].

### Liver pattern diagnoses

Philips C5-2 curvilinear transducers were used with the Lumify Application (version 4.0.1) connected to tablets with Android 9 Pie. The abdomen setting was used when acquiring ultrasound images. All images were collected in lossless digital imaging and communications in medicine (DICOM) format. Scans were performed alongside a survey programmed in Open Data Kit (version 2023.2.4) on another Android 9 Pie tablet, filled out by an assistant to the sonographer who could speak the local language and was from the district where the data collection was taking place on that day. Each participant was seen by one expert sonographer and examined within their village. Four sonographers with 5–20 years of experience on diagnosing schistosomal liver fibrosis conducted the examinations.

The data collection protocol for ultrasound images was based on the Niamey protocol, the most recent WHO guidance for staging of liver morbidity associated with *S. mansoni*. A freehand scan was undertaken, followed by acquisition of image patterns. Image patterns were assigned letters from A to F, representing different presentations of schistosomal liver fibrosis (C–F) or related fibrosis (B). B patterns were separated into three categories: B0 (feather streaks) which appears as lines across the liver; B1a/b (flying saucers or starry sky) which appears as solid white dots and small dots on the field of view; and B2 (spider thickening) which appears as thickening of many second order portal vein branches. C–F patterns, which represent periportal fibrosis (PPF), were in five categories: C1 (prominent peripheral rings) which appears as rings with no echogenic centre; C2 (prominent pipestems) which appears as brightness along a horizontal cross-section of a vessel; D (ruff) which appears as thickness around the main portal vein; E (patches) which appears as a blocked vessel which has no lumen; and F (bird’s claw) which appears as a blocked vessel with no lumen that is stretched to the periphery. There were three probe positions from which images of fibrosis patterns could be captured: the sagittal left parasternal view, a transverse view, and an oblique view of the portal vein, shown in Supplementary Figure S1. The patterns and their corresponding probe positions as provided in SchistoTrack protocol guidance are given in Supplementary Table S1.

### Model ground truth diagnoses

The liver pattern assigned to an image at the point of acquisition was used as the ground truth image-level label for model training. This label was given with context that was not present at the re-reading. This context includes participant age and sex, as well as comorbidities that were evident during the acquisition of ultrasound video sweeps and, importantly, any information from freehand scans. At the point of acquisition, the sonographers were also better aware of the precise probe position and cross-section of the liver that was in view, which better positioned them to make an informed diagnosis. Furthermore, there is evidence that radiologists perform better in the clinic than in re-reading settings [28].

The images were originally labelled in eight different classes: B0, B1, B2, C1, C2, D, E and F. For the models in this paper, the B-patterns (B0, B1(a/b) and B2) were consolidated into one class and E/F were also consolidated into one class. The B-patterns were consolidated because they all represent fibrosis that may not be caused by schistosomiasis and are diffuse patterns without any particular anatomical anchoring to the portal vessels. The E and F patterns were consolidated because there were small numbers of images for these patterns. They also represented a similar severity of PPF.

### Generating a healthy class

A healthy participant was defined as someone with no fibrosis patterns or other sonographical abnormalities (defined as having cirrhosis-like liver, fatty-like liver, hepatitis B-liver liver, polycystic kidney disease, liver cysts or situs inversus), was not pregnant, and who had fasted for at least two hours before the ultrasound examination. Images were not taken at the point of data acquisition for healthy participants since it was difficult to define what these images would be capturing, given that there was no pathology present or no clear single liver view. Instead, to generate a healthy class, four video frames were sampled from a video that swept from a good view of the gall bladder into the cephalic position from 100 randomly selected healthy participants. The four video frames were sampled as follows: two from approximately one-third of the way through the video, and two from two-thirds of the way through. This sampling was done to give examples of two different transverse cross-sections of the liver that should be in the same view as the B, C2, E and F patterns. The video frames were processed in the same way as the images; however, there were no labels on these frames.

### Train and test splits

Train and test splits were assigned at a ratio of 90:10 such that there was representation of each class in the test set. 10-fold cross validation was carried out to assess the performance of each model setup. Folds were stratified on a participant level based on the highest pattern of fibrosis present. Participants in the healthy class were randomly assigned to the train and test sets in a ratio of 90:10, and those in the train set were randomly assigned to each fold during cross validation. Splits were assigned on a participant level to avoid data leakage.

### Study participants

The total number of participants in 2023 with sonography survey data was 3186, and the number who also had ultrasound imaging data was 3095. The total number of participants in 2024 with sonography survey data was 3311, and the number who also had ultrasound imaging data was 3242. The number of participants who had ultrasound images of liver fibrosis patterns in either 2023 or 2024 was 1433, with a total of 3710 images of fibrosis patterns being collected across the two years. Therefore, the total number of participants that were included in the analysis was 1533, inclusive of the 100 healthy participants who contributed video frames. 377 participants contributed images in both years. The only exclusion criterion for participants with liver fibrosis patterns was the presence of the B (unclear) pattern, which resulted in 14 participants being excluded. Aside from this criterion, all participants with ultrasound images of fibrosis patterns were included. Therefore, there were no exclusion criteria based on age, sex, fasting or any other abnormal sonographical findings in the course of the examination. This was done to reflect that an image classification tool should be as generalisable to as many subjects as possible. Moreover, excluding participants based on other abnormalities would have excluded many severe cases. For example, participants with patterns E/F also disproportionately had abnormalities such as oesophageal varices, therefore excluding individuals with abnormalities would have further biased the dataset in favour of the less severe patterns.

### Preprocessing

For this analysis, the pixel array was extracted from the digital imaging and communications in medicine (DICOM) format images and used for image-level classification. At the point of image acquisition, the fibrosis pattern label was written on the image, outside of the field of view, using the Lumify application. Other text, such as the participant barcode and information on the Lumify display, also appeared outside of the field of view. The text was removed in the preprocessing step by using OpenCV [32] to find the contours in the image, identify the largest contour (which was always the ultrasound field of view), and preserve the pixels inside this contour while setting all other pixels to zero.

Data augmentations were used to enlarge the dataset and to mitigate the effects of heterogeneous classes. These included changes to the pixel values of the image, including small, random changes of contrast, brightness and saturation, as well as Gaussian blur. Geometric transformations were also applied, such as random horizontal flip, cropping and resizing, and random affine transformations such as rescaling and translation. These transformations were implemented using the torchvision package (version 0.16.0) [33].

### Models

Python 3.10 was used for all analyses. The setup was a 6-class classification problem, comprising the five fibrosis pattern classes (B, C1, C2, D and E/F), and the healthy class. SchistoTrackNet and SchistoTrackNet-Random were CNN-based models. SchistoTrackNet was trained on liver ultrasound image data from SchistoTrack using supervised contrastive (SupCon) loss [34] and was initialised using PULSENet weights. The variant of PULSENet used was trained to classify 13 views of fetal anatomy using the PULSE dataset, a large fetal ultrasound dataset [15, 16]. PULSENet was used as an ultrasound world encoder, and despite being trained on fetal ultrasound images rather than liver ultrasound images, encode features relevant to ultrasound imaging. The architecture of PULSENet was based on VGG-16 [35]. SchistoTrackNet-Random was trained in the same manner as SchistoTrackNet using SupCon loss, but had random weight initialisation. SupCon loss takes an ‘anchor’ image, in addition to many positive examples (images from the same class as the anchor) and negative examples (images from any other class), encouraging normalised feature embeddings of the anchor and the positive samples to be pulled together, while encouraging the feature embeddings of the anchor and negative samples to be pulled apart. For the transformer-based models, ViT-base was used and weights were initialised using pre-training from the ImageNet dataset [17]. Three loss functions were compared: SupCon loss (ViT-SupCon), focal loss (ViT-Focal, to account for class imbalance), and cross entropy loss (ViT-CE). Each model was trained for 200 epochs, with the epoch that gained the highest classification accuracy on the holdout set for each fold being chosen to evaluate the model.

For SchistoTrackNet and SchistoTrackNet-Random, two adaptation layers were used as the classification head. These were 1 × 1 convolutional layers, that localise the relevant information to a high spatial resolution [36]. A linear layer was used for the transformer-based models. In the cases where SupCon loss was used, weights for the backbone encoder were frozen and the classifier was trained separately. The classification head was trained using focal loss. Probabilities for each class were computed by applying the sigmoid function to the classification head outputs in the case of the linear layer. In the case of the adaptation layers, the outputs were already between 0 and 1. Final classifications were determined by applying a softmax layer to these probabilities.

The following hyperparameters were tuned: SupCon loss temperature, focal loss gamma, and learning rate, for scenarios outlined in Supplementary Table S2. The temperature parameter in SupCon loss has been shown to control the strength of penalties on the hard negative samples, with lower values of temperature increasing the penalty on hard-to-classify samples [37]. The focal gamma parameter in focal loss controls the extent to which easy-to-learn samples (ones that are in classes with many examples) are downweighted. Therefore, higher values of gamma force the model to focus on correctly classifying harder examples. The learning rate determines the step size for each iteration during the optimisation of the model weights. Bayesian hyperparameter tuning with 10 iterations was used to select values for these hyperparameters, optimising for classification accuracy. Adam was used as the optimiser for all experiments.

As a sensitivity analysis, the classification of the available fibrosis pattern images without a healthy class (a 5-class problem), was performed.

### Selected hyperparameter values

For SchistoTrackNet and SchistoTrackNet-Random, the learning rate was tuned to 2.18 *×* 10^*−*4^, while the temperature was tuned to 0.24. For ViT-SupCon, the learning rate was tuned to 9.69 *×* 10^−6^, and the temperature was tuned to 1.19. For ViT-focal, the learning rate was tuned to 4.99 *×* 10^−6^, and focal gamma was tuned to 4.

### Performance metrics

A number of metrics and visualisation techniques were used to evaluate models and understand their performance. For visualisation, uniform manifold approximation and projection (UMAP) was used. UMAP is a dimensionality reduction technique used to interpret high-dimensional data, in particular to see if there are clusterings within these data. For model evaluation, accuracy and F1-score were used in addition to balanced accuracy, which is defined as the arithmetic mean of sensitivity and specificity. Confidence interval estimates for metrics were obtained by taking 1000 bootstrap samples of the predictions made by each model. The metrics were calculated for each of these samples and the 2.5th and 97.5th percentiles were taken to construct 95% confidence intervals [38]. 10-fold cross validation with stratified splits was used to evaluate model performance, with Friedman’s *χ*^2^-test being used to test for any difference in F1-score between the five models with post-hoc Wilcoxon signed-rank tests with a Holm correction run for differences between pairs of models if the *χ*^2^-test was significant. PPVs and NPVs were calculated for each class using the predictions given on the test set when the softmax function was applied to the probabilities output by the classification head of each model, though they may be inflated for some liver patterns because they were not calculated using a fixed sensitivity threshold. For SchistoTrackNet, activation maps were produced using GRAD-CAM [18]. For ViT-SupCon, attention rollout was used to generate attention maps to visualise the area of the image important to the classification decision [20].

### Reading of images by a second sonographer

At 2023 data collection, image-level labels were given to the ultrasound images of fibrosis patterns at the point of acquisition, only allowing for one fibrosis pattern to be assigned per image. Each image was re-read by a second sonographer in Kampala two to three months after participant examination in the field, at which point multiple patterns were allowed to be assigned per image. The second readers were the same set of sonographers that collected ultrasound images in the field, but the participants were swapped so that one sonographer re-read all the images that were acquired by another sonographer. The only information the second reader had available was the ultrasound image itself and that a fibrosis pattern was assigned in the field since there were no images taken of healthy participants. There was no participant information available. In 2024, the field procedure and second reader procedure remained the same, except that second readers were randomly allocated images acquired by other sonographers rather than reading all images from one other sonographer. Agreement between the sonographer is shown in Figure 5.

**Fig. 5:**
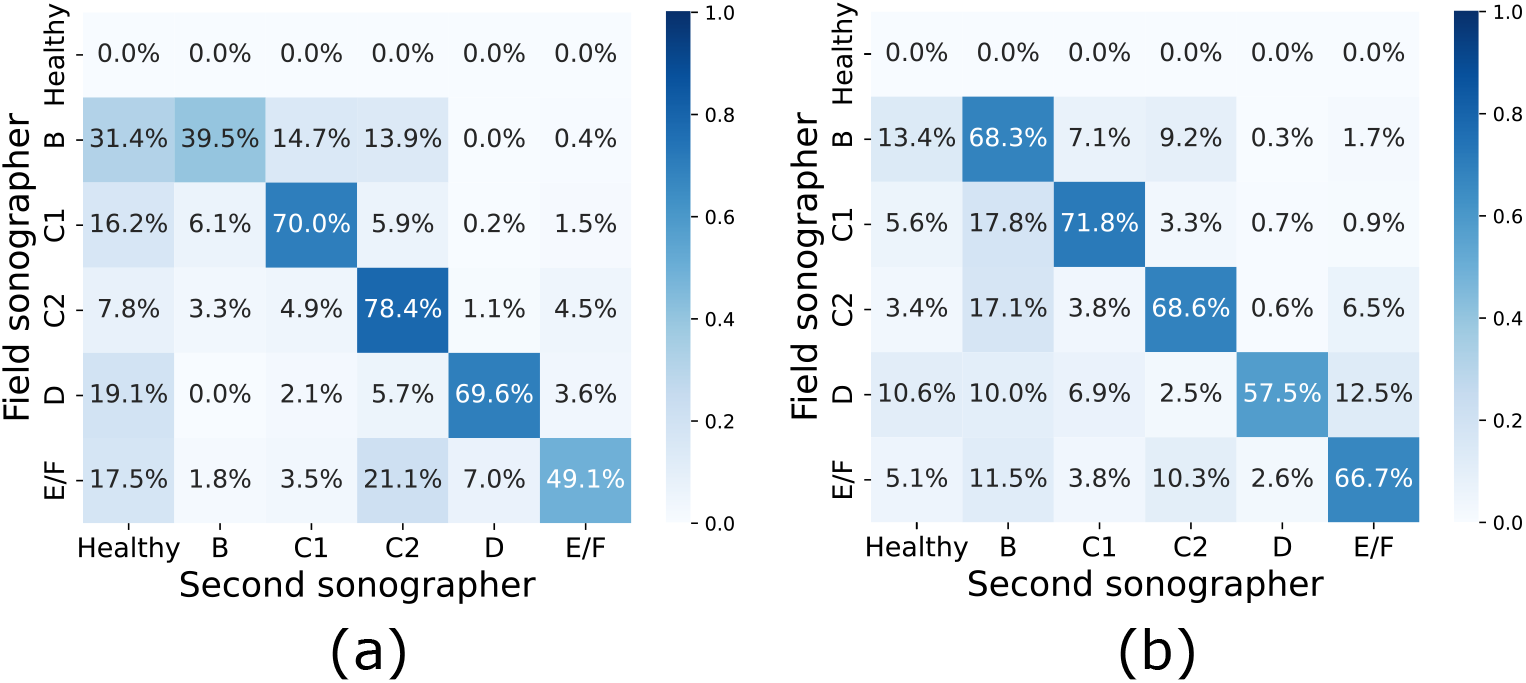
Sonographer agreement. Confusion matrices showing the sonographer agreement among the curated images for (a) 2023 and (b) 2024.

## Data availability

Image data are protected and are not available due to data privacy laws, restrictions in ethics approvals and participant agreements associated with the ongoing SchistoTrack Cohort study. These restrictions prevent open sharing of raw images and individual-level participant information that may identify individuals.

## Code availability

Code has been provided as supplementary material.

## Acknowledgments

We are thankful for the involvement of our study participants and the SchistoTrack teams, especially the sonographers, nurses and surveyors. We would also like to thank the Uganda Ministry of Health, local district leaders, focal health workers, and village health teams. We are grateful for the constant involvement through the study time-points and community engagement meetings of our study participants. Special thanks also to the SchistoTrack and Noble Groups in Oxford for everyday discussions and feedback, and for the feedback from Bartek Papiez.

## Author contributions

Conceptualisation: EO, AN, GFC. Data curation: EO, VA, SM, TM, BNt, BNa, and GFC. Formal analysis: EO. Funding acquisition: EO and GFC. Investigation and Methodology: EO, AN, and GFC. Project administration: BNa, NBK, and GFC. Resources: GFC. Software: GFC. Supervision: AN and GFC. Validation: EO, VA, SM, TM, BNt, and GFC. Visualisation: EO. Writing - original draft: EO. Writing - review & editing: EO, VA, SM, TM, BNt, BNa, NBK, AN and GFC.

## Ethics approvals

Data collection and use were reviewed and approved by Oxford Tropical Research Ethics Committee (OxTREC 509-21), Vector Control Division Research Ethics Committee of the Uganda Ministry of Health (VCDREC146), and Uganda National Council of Science and Technology (UNCST HS 1664ES). Written informed consent was obtained from adult participants aged 18 years and older, who also provided written consent on behalf of verbally assented children with fingerprint assent. Older children also provided written consent in addition to the adult consent on their behalf.

