## Supplements for "SchistoTrackNet: machine learning for diagnosis of schistosomiasis-associated periportal fibrosis from ultrasound images"

### Supplementary Information S

#### S.1 Probe positions

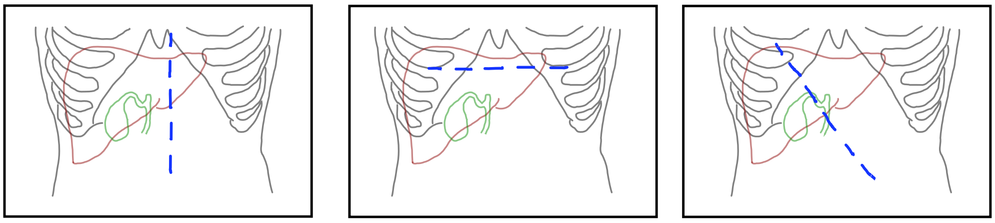

(a) (b) (c)

**Fig. S1**: **Views of the liver.** Diagrams to show the views of the liver where fibrosis patterns are found, based on the Niamey protocol [39].

**Table S1**: Niamey protocol patterns and their respective probe position association instructed by SchistoTrack.

| Niamey protocol pattern | Probe angle |
|---|---|
| B | Transverse |
| C1 | Sagittal |
| C2 | Transverse |
| D | Oblique |
| E | Transverse |
| F | Transverse |

#### S.2 Hyperparameter tuning

**Table S2**: **Hyperparameter tuning.** An outline of the different configurations of the model training that required different hyperparameters to be tuned.

| Run | Backbone | Loss | Classifier | Hyperparameters to tune |
|---|---|---|---|---|
| 1 | SonoNet | Contrastive | Adaptation layers | Temperature, learning rate |
| 2 | ViT | Contrastive | Linear layer | Temperature, learning rate |
| 3 | ViT | Focal | Linear layer | Focal gamma, learning rate |

#### S.3 Participant characteristics

**Table S3**: **Number of images for each fibrosis pattern.** This table shows the number of images of each liver fibrosis pattern were in the train and test sets, split by year. Train and test splits were done at a participant level to ensure no data leakage. Entries are presented as number of images (number of participants), where the number of images is larger since participants could contribute multiple images of the same liver fibrosis pattern.

| Pattern | Train | | Test | | Overall |
|---|---|---|---|---|---|
| | 2023 | 2024 | 2023 | 2024 | |
| B | 412 (336) | 677 (535) | 56 (47) | 70 (54) | 1215 (848) |
| C1 | 414 (392) | 507 (489) | 43 (41) | 67 (65) | 1031 (789) |
| C2 | 406 (389) | 466 (448) | 43 (40) | 60 (59) | 975 (748) |
| D | 174 (161) | 145 (139) | 20 (17) | 15 (14) | 354 (260) |
| E/F | 50 (36) | 66 (52) | 7 (5) | 12 (7) | 135 (84) |
| **Total** | | | | | 3710 (1433) |

**Table S4**: Distributions of liver fibrosis patterns among adults and children. Adults were defined as participants aged $\geq 15$ and children were participants aged <15.

| | 2023 | | 2024 | |
|---|---|---|---|---|
| | Children | Adults | Children | Adults |
| B0 | 11.2% (151/1345) | 4.8% (88/1841) | 12.0% (157/1310) | 7.2% (145/2001) |
| B1a | 10.0% (134/1345) | 3.3% (61/1841) | 12.1% (159/1310) | 5.7% (115/2001) |
| B1b | | | 1.9% (25/1310) | 2.2% (44/) |
| B2 | 1.0% (14/1345) | 2.1% (39/1841) | 0.9% (12/1310) | 3.8% (77/2001) |
| C1 | 6.1% (82/1345) | 20.6% (380/1841) | 7.9% (104/1310) | 23.2% (465/2001) |
| C2 | 6.2% (84/1345) | 20.2% (372/1841) | 7.3 (96/1310) | 21.7% (435/2001) |
| D | 0.4% (5/1345) | 9.8% (180/1841) | 0.3% (4/1310) | 7.7% (155/2001) |
| E | 0% (0/1345) | 2.4% (45/1841) | 0.1% (1/1310) | 3.0% (60/2001) |
| F | 0% (0/1345) | 0.4% (8/1841) | 0% (0/1310) | 0.6% (13/2001) |

**Table S5**: **Distribution of fibrosis pattern images among children and adults.**

| Pattern | 2023 | | 2024 | |
|---|---|---|---|---|
| | Children | Adults | Children | Adults |
| B | 299 | 188 | 353 | 381 |
| C1 | 82 | 380 | 104 | 465 |
| C2 | 84 | 372 | 96 | 435 |
| D | 5 | 180 | 4 | 155 |
| E/F | 0 | 53 | 1 | 73 |

#### S.4 UMAP projections of feature vectors

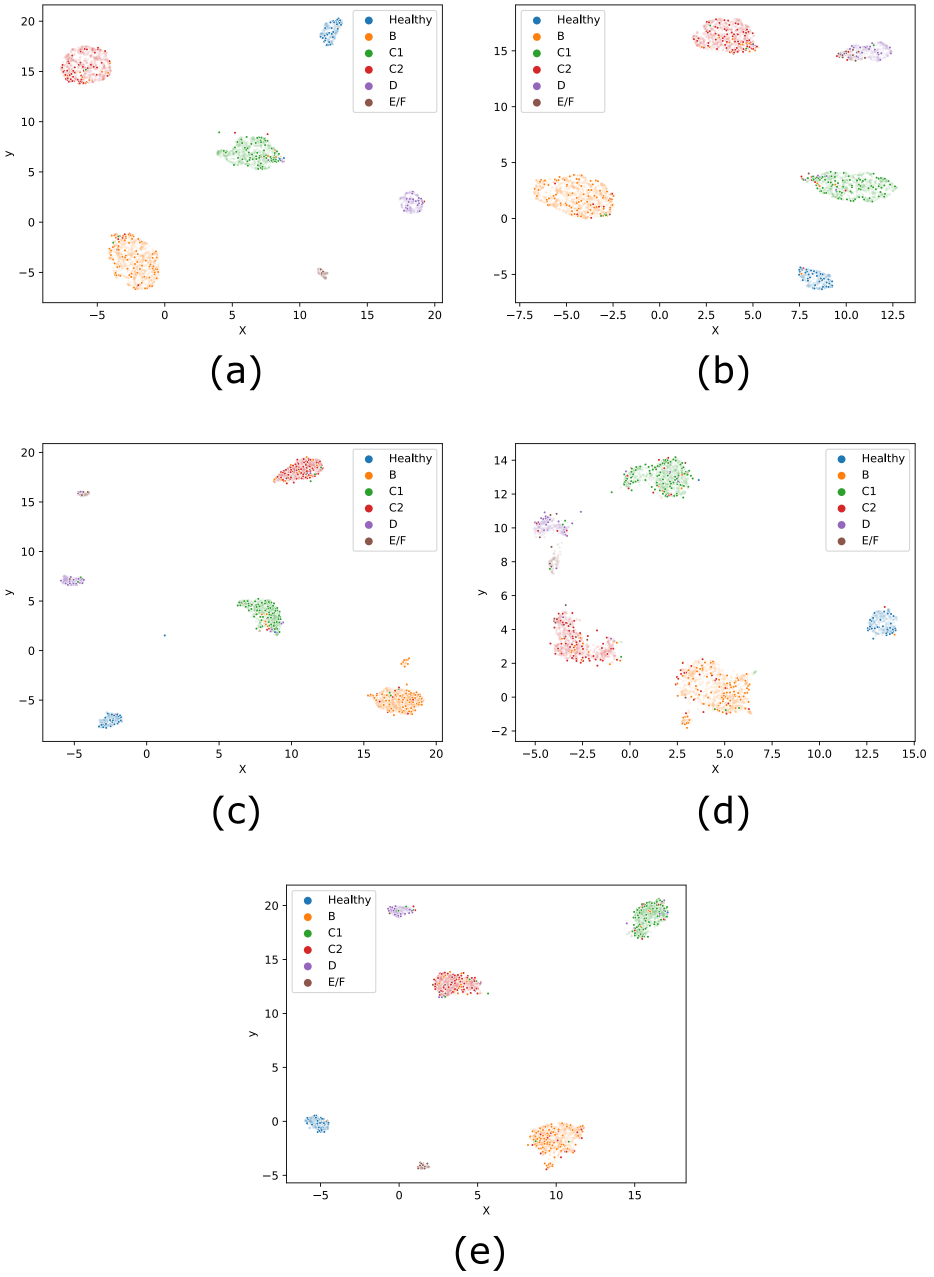


**Fig. S2**: **UMAP projections for models with a healthy class.** Projections of feature vectors from the test set of curated images and healthy video frames to 2D space. (a) SchistoTrackNet. (b) SchistoTrackNet-Random. (c) ViT-SupCon. (d) ViT-Focal. (e) ViT-CE.

#### S.5 Results split by year

**Table S6**: **Quantitative results by year.** A summary of the quantitative results gained from the different models, split by year.

| Model (loss) | Accuracy | Balanced accuracy | F1-score |
|---|---|---|---|
| *2023* | | | |
| *CNN-based* | | | |
| SchistoTrackNet | 0.8517 | 0.8517 | 0.8541 |
| SchistoTrackNet-Random | 0.8230 | 0.6981 | 0.8108 |
| *Transformer-based* | | | |
| ViT-SupCon | 0.8278 | 0.7282 | 0.8205 |
| ViT-Focal | 0.7895 | 0.7278 | 0.7853 |
| ViT-CE | 0.8038 | 0.7306 | 0.8009 |
| *2024* | | | |
| *CNN-based* | | | |
| SchistoTrackNet | 0.7946 | 0.7487 | 0.8014 |
| SchistoTrack-Random | 0.7857 | 0.6388 | 0.7695 |
| *Transformer-based* | | | |
| ViT-SupCon | 0.7768 | 0.6675 | 0.7774 |
| ViT-Focal | 0.7813 | 0.6645 | 0.7736 |
| ViT-CE | 0.7545 | 0.6789 | 0.7561 |

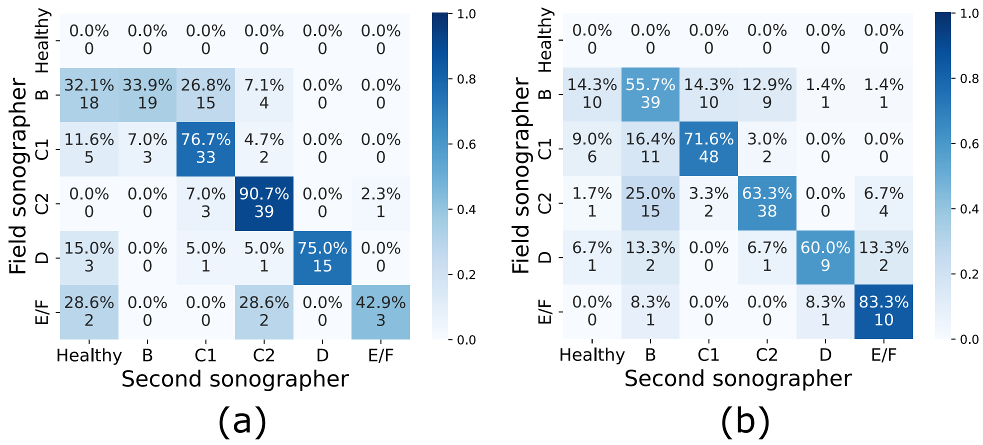


**Fig. S3**: **Sonographer agreement by year.** (a) 2023. (b) 2024.

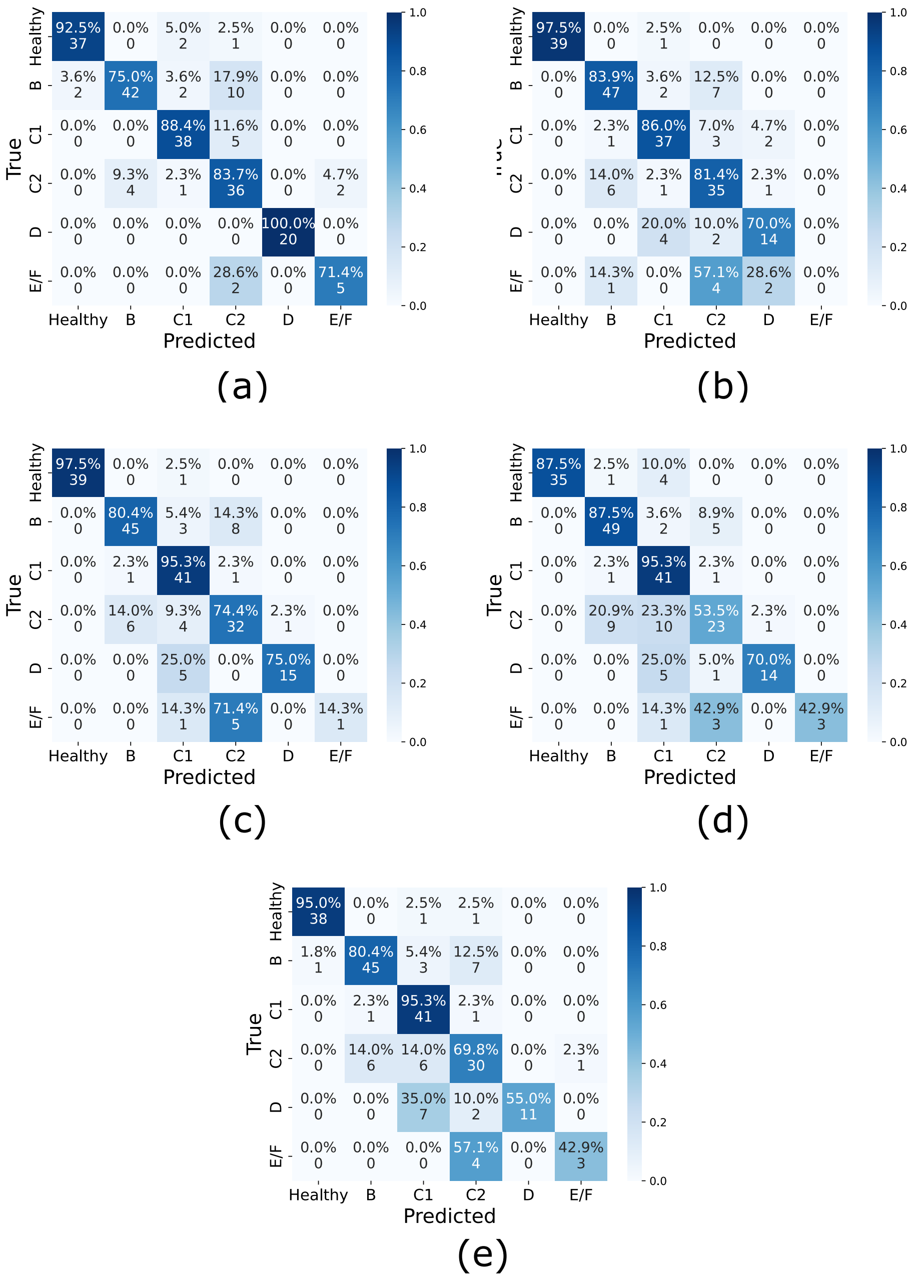


**Fig. S4**: **Confusion matrices for deep learning-based models, 2023 images only.** (a) SchistoTrackNet. (b) SchistoTrackNet-Random. (c) ViT-SupCon. (d) ViT-Focal. (e) ViT-CE.

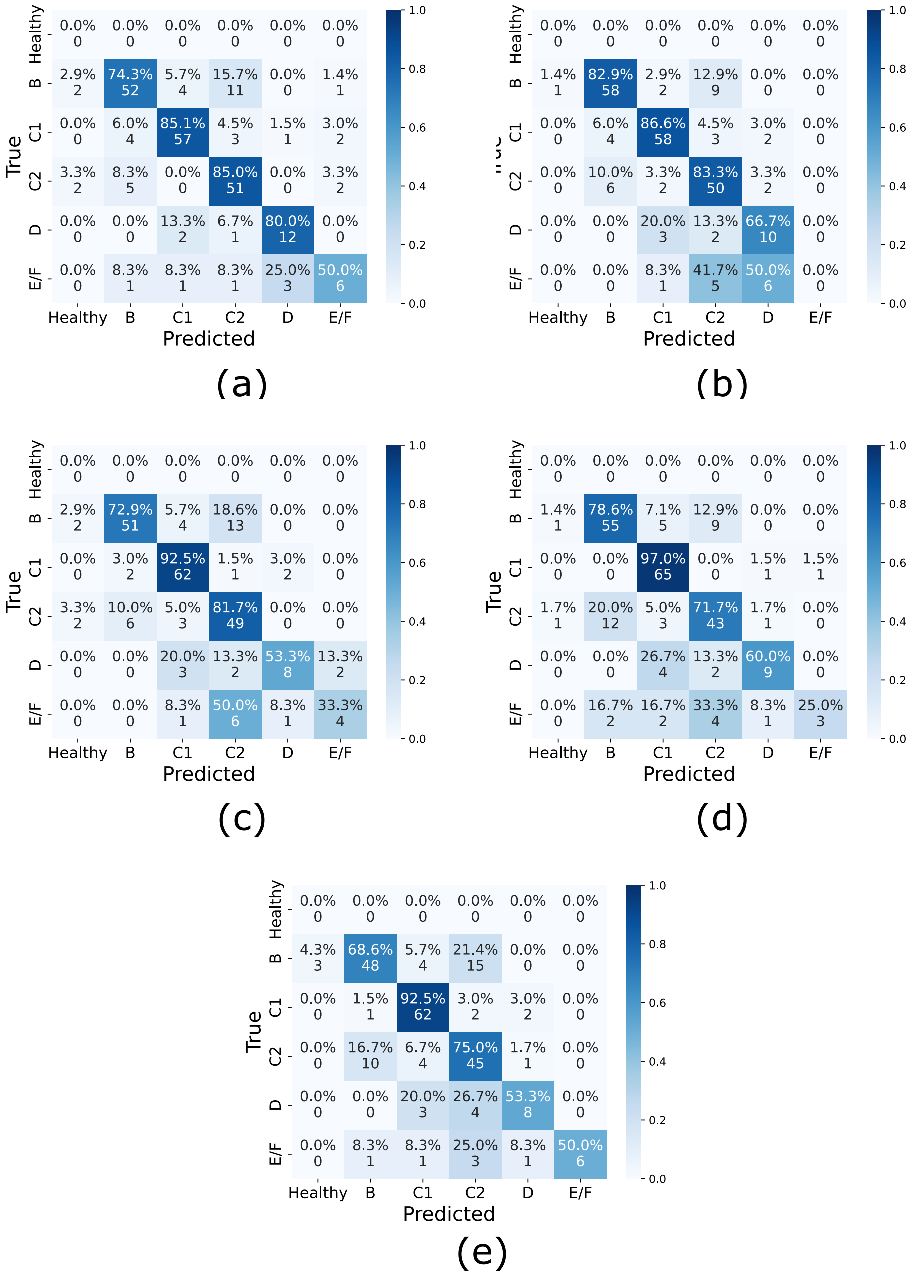


**Fig. S5**: **Confusion matrices for deep learning-based models, 2024 images only.** (a) SchistoTrackNet. (b) SchistoTrackNet-Random. (c) ViT-SupCon. (d) ViT-Focal. (e) ViT-CE.

#### S.6 Confusion matrices by age and sex

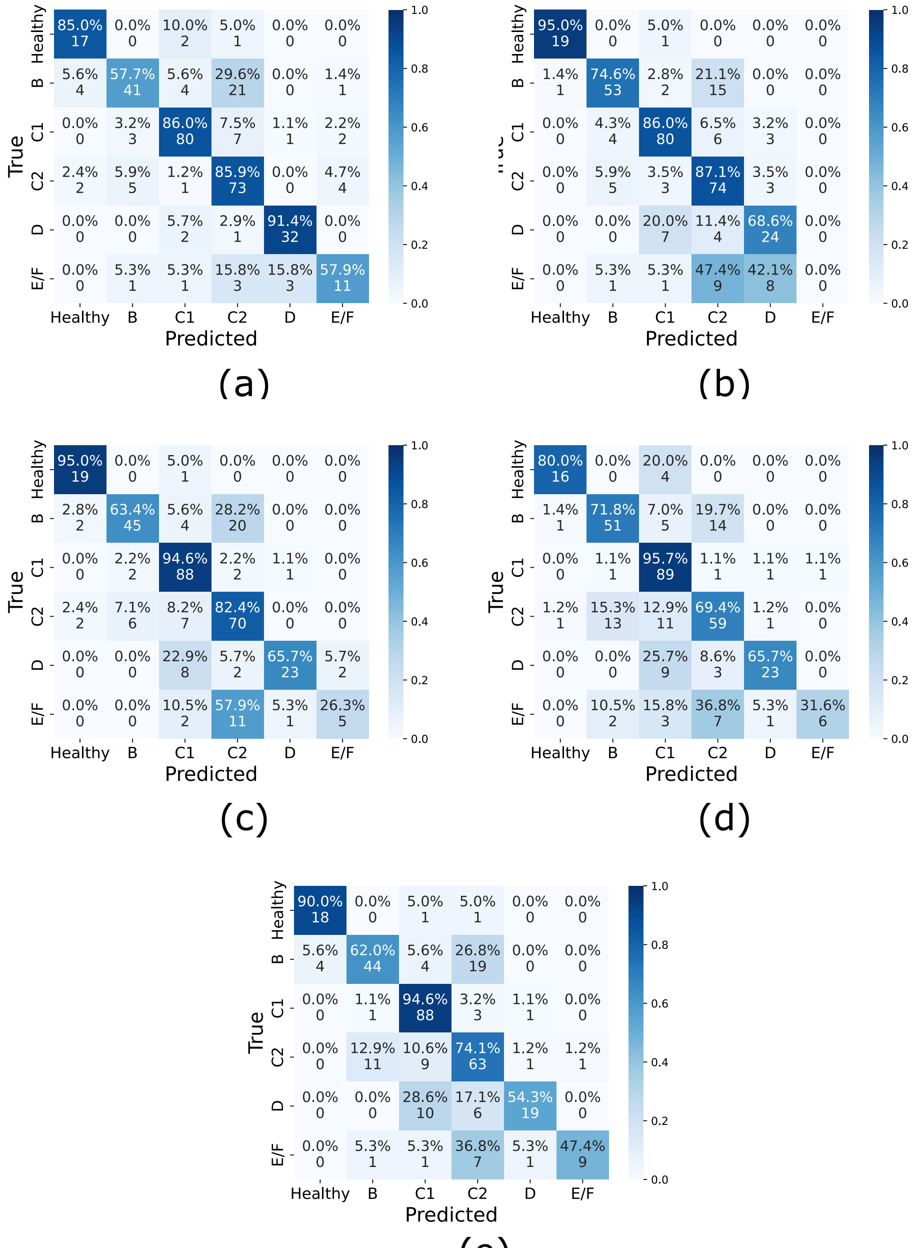


**Fig. S6**: **Confusion matrices for deep learning-based models, adult participants only.** Adults were defined as aged $\geq$ 15. (a) SchistoTrackNet. (b) SchistoTrackNet-Random. (c) ViT-SupCon. (d) ViT-Focal. (e) ViT-CE.

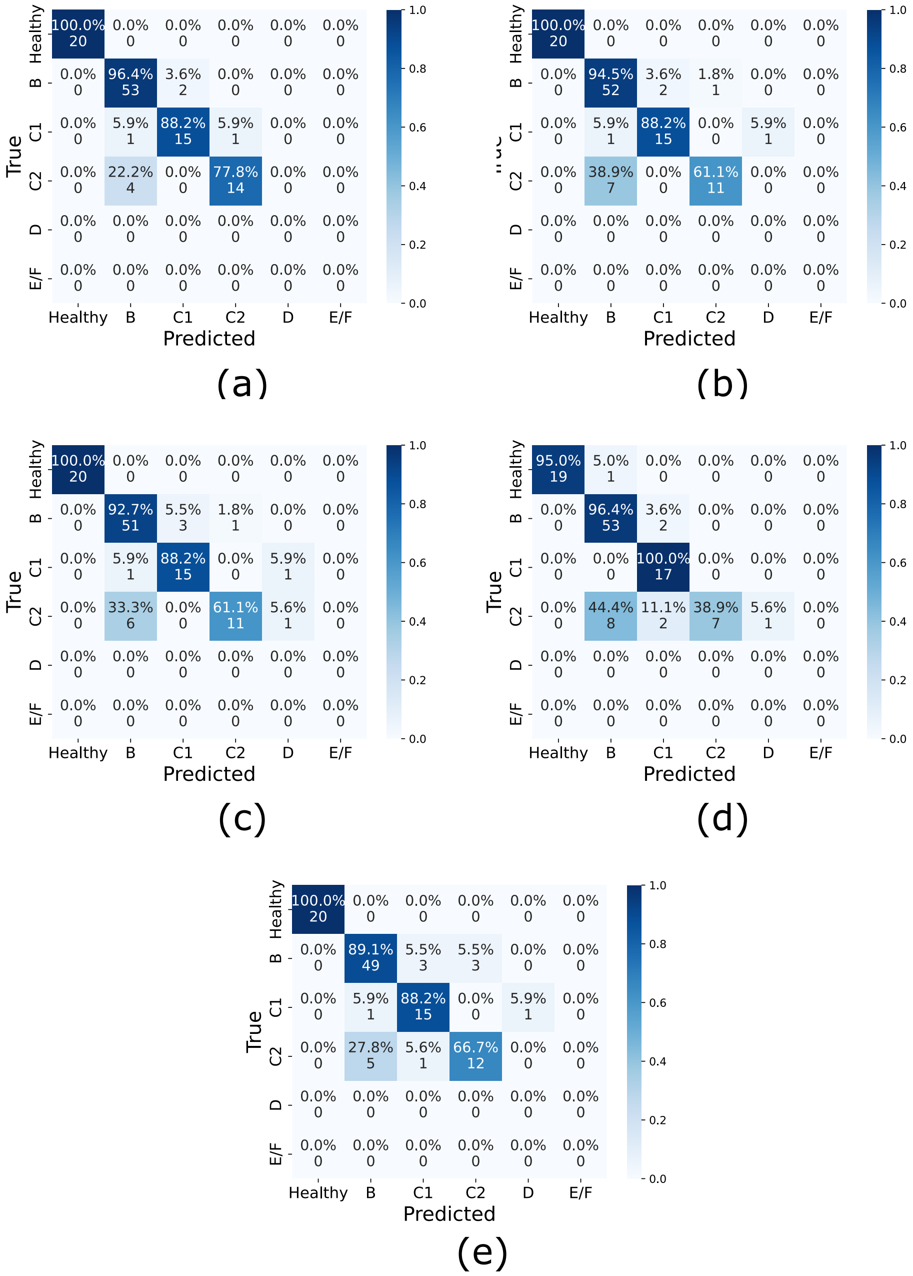


**Fig. S7**: **Confusion matrices for deep learning-based models, child participants only.** Children were defined as aged $< 15$. (a) SchistoTrackNet. (b) SchistoTrackNet-Random. (c) ViT-SupCon. (d) ViT-Focal. (e) ViT-CE.

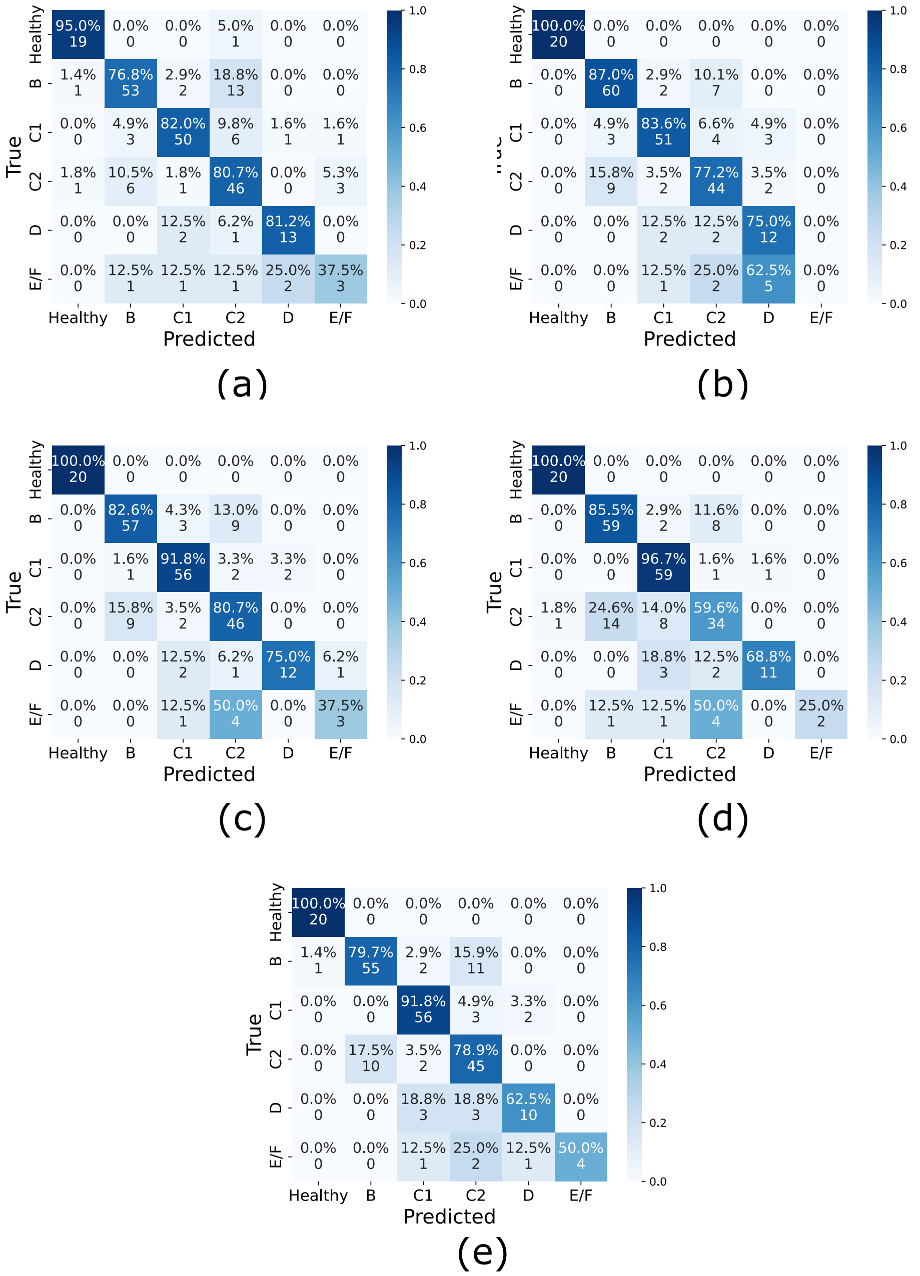


**Fig. S8**: **Confusion matrices for deep learning-based models, female participants only.** (a) SchistoTrackNet. (b) SchistoTrackNet-Random. (c) ViT-SupCon. (d) ViT-Focal. (e) ViT-CE.

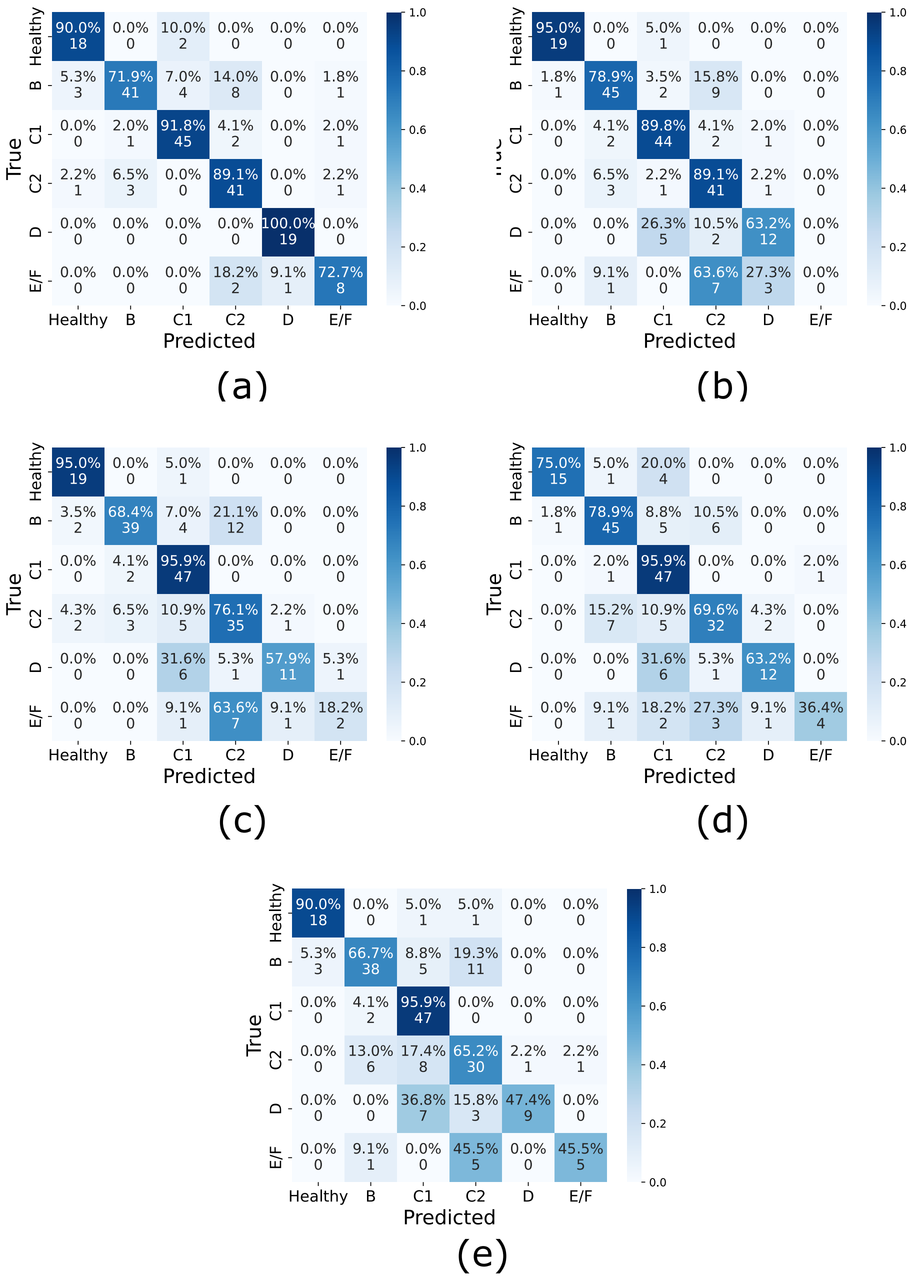


**Fig. S9**: **Confusion matrices for deep learning-based models, male participants only.** (a) SchistoTrackNet. (b) SchistoTrackNet-Random. (c) ViT-SupCon. (d) ViT-Focal. (e) ViT-CE.

#### S.7 Curated image models

**Table S7: Quantitative results.** A summary of the quantitative results gained from the different models that were used, for curated images without a healthy class. Bootstrapped confidence intervals are given in square brackets.

| **Model (loss)** | **Accuracy** | **Balanced accuracy** | **F1-score** | **Any agreement** |
|---|---|---|---|---|
| *CNN-based* | | | | |
| SchistoTrackNet | 0.7761 [0.733, 0.817] | 0.6823 [0.623, 0.739] | 0.7714 [0.726, 0.815] | 0.8244 |
| *Transformer-based* | | | | |
| ViT-SupCon | 0.7786 [0.738, 0.819] | 0.7136 [0.653, 0.776] | 0.7804 [0.740, 0.821] | 0.8295 |
| ViT-Focal | 0.7761 [0.730, 0.814] | 0.6942 [0.638, 0.752] | 0.7711 [0.724, 0.811] | 0.8219 |

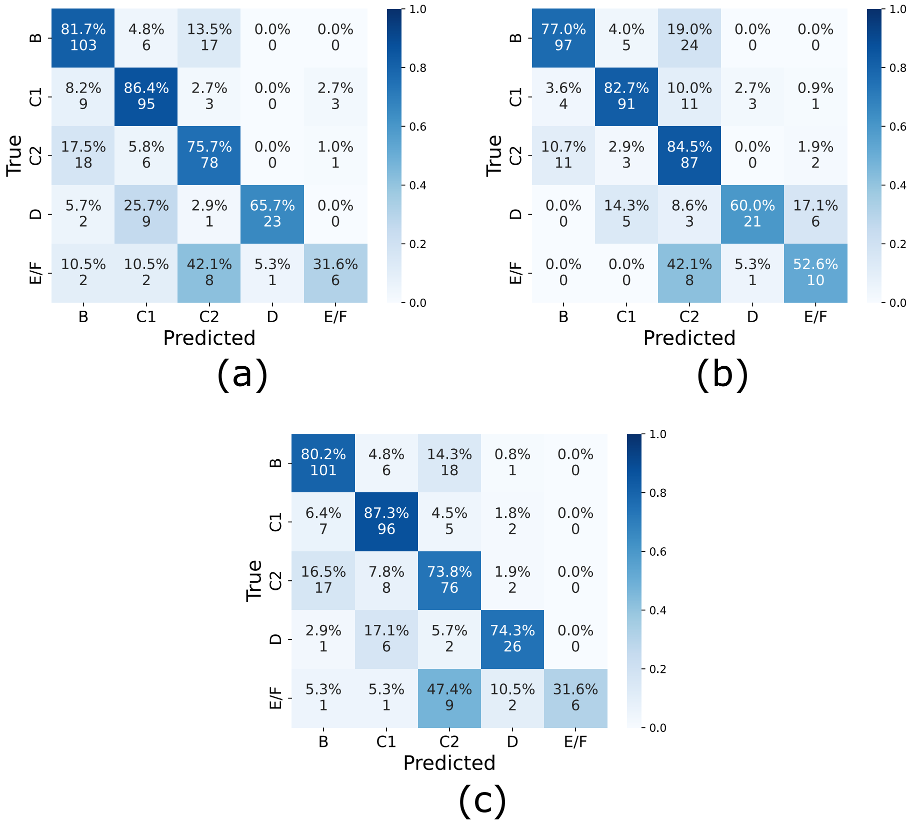


**Fig. S10**: **Confusion matrices for image-only model.** Confusion matrices for the curated images only for each model setup for the test set. (a) SchistoTrackNet. (b) ViT-SupCon. (c) ViT-Focal.

#### S.8 Attention maps

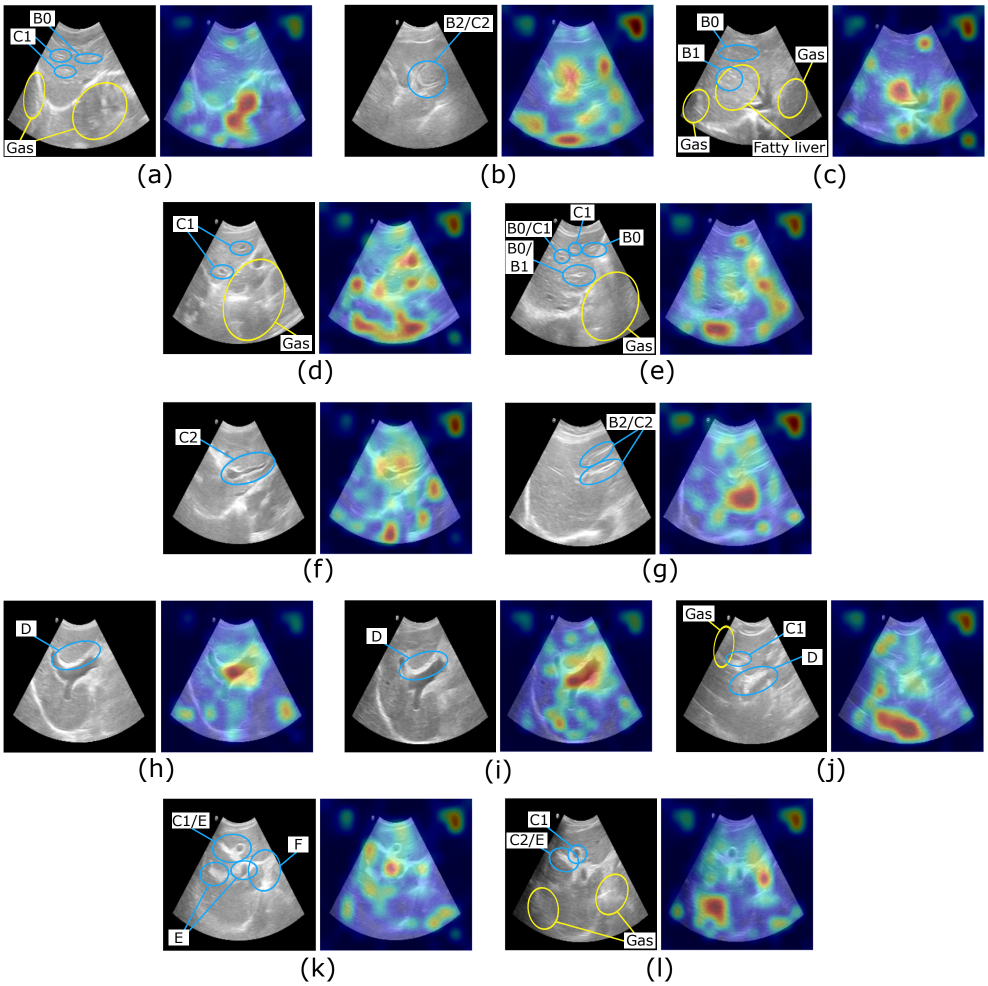


**Fig. S11**: **Annotated ultrasound images with attention maps.** Ultrasound images for each grade and with different predictions (correctly or incorrectly predicted by ViT-SupCon) alongside attention maps given by attention rollout from ViT-SupCon. (a) B0, correctly predicted. (b) B2, incorrectly predicted (C2). (c) B1b, incorrectly predicted (C1). (d) C1, correctly predicted. (e) C1, incorrectly predicted (B). (f) C2, correctly predicted. (g) C2, incorrectly predicted (B). (h) D, correctly predicted. (i) D, correctly predicted. (j) D, incorrectly predicted (C1). (k) E, correctly predicted. (l) E, incorrectly predicted (C2).
